# CHAPAS-4 trial: second-line anchor drugs for children with HIV in Africa

**DOI:** 10.1101/2024.04.12.24305333

**Authors:** Mutsa Bwakura-Dangarembizi, Alexander J Szubert, Vivian Mumbiro, Cissy M Kityo, Abbas Lugemwa, Katja Doerholt, Chishala Chabala, Mary Nyathi, Bwendo Nduna, David Burger, Clare Shakeshaft, Kusum Nathoo, Victor Musiime, Ibrahim Yawe, Annabelle South, Joyce Lungu, Wedu Ndebele, Mwate Mwamabazi, Anna Griffiths, Rashidah Nazzinda, Kevin Zimba, Yingying Zhang, Simon Walker, Anna Turkova, A Sarah Walker, Alasdair Bamford, Diana M Gibb, CHAPAS-4 Trial Team

## Abstract

**Background:** Children living with HIV requiring second-line antiretroviral therapy (ART) have limited options, an unmet need considering children require life-long ART.

**Methods:** Children from Uganda, Zambia, Zimbabwe were randomised to one of four second-line anchor drugs: dolutegravir(DTG), ritonavir-boosted darunavir(DRV/r), atazanavir(ATV/r), or lopinavir(LPV/r) in the factorial CHAPAS-4 trial (second randomisation to tenofovir alafenamide fumarate(TAF) or standard-of-care(SOC) backbone, reported elsewhere). Dosing followed WHO weight-bands. The primary endpoint was viral load(VL) <400copies/mL at week-96, analysed using logistic regression, hypothesising that DTG and DRV/r would be superior (threshold p=0.03) to LPV/r and ATV/r arms combined and ATV/r would be non-inferior to LPV/r(12% margin). Secondary endpoints included immunology and safety. Analyses were intention-to-treat.

**Results:** 919 children, median(IQR) age 10(8-13) years, 54% male, baseline VL 17,573(5549,55700) copies/mL, CD4 669(413, 971) cells/mm^3^, weight-for-age Z-score -1.6(-2.4,-0.9), had spent median(IQR) 5.6(3.3,7.8) years on first-line ART. At week-96, DTG was superior (by 9.7%(95% CI 4.8%, 14.5%); p<0.0001) and DRV/r showed a trend to superiority(by 5.6%(0.3%, 11.0%); p=0.04) compared to LPV/r and ATV/r arms combined. ATV/r was non-inferior to LPV/r(+3.4%(-3.4%,+10.2%); p=0.33). CD4 counts increased with no differences between arms. Toxicity was lowest with DTG. All arms except LPV/r showed age-appropriate weight/height gains at week-96. DTG was not associated with excess absolute weight-gain(<1kg) vs. DRV/r or ATZ/r, irrespective of backbone randomisation.

**Conclusions:** DTG-based regimens are safe and cost-effective for second-line ART. DRV/r and ATV/r are also good options. Fixed-dose combinations of DTG, DRV/r or ATV/r with nucleoside/nucleotide-reverse-transcriptase-inhibitors(NRTIs) would increase access to robust, essential second-line options for children.(ISRCTN22964075)

## Background

Globally, numbers of children living with HIV (CLHIV) accessing first-line antiretroviral therapy (ART) have increased; coupled with increasing HIV viral load (VL) monitoring, numbers requiring second-line and subsequent ART following virological failure will also increase.^1–3^ The majority of CLHIV live in Africa and until recently first-line non-nucleoside-reverse-transcriptase (NNRTI)-containing regimens were most frequently used.^3^ For second-line ART following NNRTI failure, guidelines recommend an anchor drug from a new class (boosted protease inhibitor (bPI) or integrase inhibitor (INSTI)), combined with 2 nucleoside-reverse-transcriptase-inhibitors (NRTIs). Maximising effectiveness and tolerability while minimising toxicity is particularly important for children who need life-long ART.^4^ The question of which anchor drugs are most effective in second-line ART for children remains unanswered.

Whereas dolutegravir (DTG) is available in child-friendly formulations, bPIs, although providing sustainable VL suppression and high barrier to resistance,^5^ have paediatric formulation challenges.^6^ Lopinavir (LPV) is currently the only paediatric ritonavir co-formulated bPI, but requires twice-daily dosing and has poor palatability; ritonavir-boosted darunavir (DRV/r) and atazanavir (ATV/r) are dosed once-daily but are not widely available as fixed-dose combinations (FDCs) for children and DRV/r is relatively costly.

The CHAPAS-4 trial compared the efficacy, safety, tolerability and pharmacokinetics of DRV/r, ATV/r, LPV/r and DTG in African children aged 3-15 years.

## Methods

CHAPAS-4 (ISRCTN22964075) was a randomised, open-label trial with a factorial design (2X4). The trial was approved by ethics committees in Uganda, Zambia, Zimbabwe, and United Kingdom. The protocol is available at www.mrcctu.ucl.ac.uk/studies/all-studies/c/chapas-4. Participants were recruited at six clinical centres in three sub-Saharan African countries: Uganda (Joint Clinical Research Centre (JCRC), Kampala; JCRC, Mbarara), Zambia (University Teaching Hospital, Lusaka; Arthur Davison Children’s Hospital, Ndola) and Zimbabwe (University of Zimbabwe Clinical Research Centre, Harare; Mpilo Central Hospital, Bulawayo).

Participants were CLHIV aged 3-15 years, weighing ≥14kg, requiring second-line ART for virologic failure defined as VL>1000 copies/ml with or without immunological and/or clinical failure. Children had to be able to swallow tablets, and post-menarchal females required a negative pregnancy test. Guardians provided written informed consent, with additional assent from older children, according to national guidelines. Children were excluded if they had severe hepatic impairment (alanine aminotransferase (ALT) ≥5 times upper-limit of normal (ULN), or ALT ≥3xULN and bilirubin ≥2xULN or clinical liver disease).

Participants were randomised to one of four anchor drugs (DTG, DRV/r, ATV/r or LPV/r) and simultaneously to one of two backbones (tenofovir alafenamide fumarate(TAF)/emtricitabine(FTC) or standard-of-care(SOC) (abacavir(ABC)/lamivudine(3TC) or zidovudine (ZDV)/3TC, whichever was not used first-line). Randomisation was stratified by centre and first-line NRTI (ABC or ZDV). A computer-generated sequential randomisation list with variably sized permuted blocks was prepared by the trial statistician and incorporated securely into an online database. The allocation was concealed until eligibility was confirmed by local centre staff, who then performed the randomisation.

Participants were seen at screening, ART switch (week 0), 2, 6, 12 weeks and 12 weekly thereafter to at least week-96 (primary endpoint): extended follow-up continued through 2 February 2023. Children with tuberculosis at enrolment or during follow-up had TAF/FTC and anchor drugs adjusted during anti-tuberculosis treatment to account for rifampicin drug-drug interaction. Additional measures ensured participant follow up during the COVID-19 pandemic (Panel 1 Supplementary Appendix 1).

The primary outcome was percentage of children alive with VL <400 copies/ml at week 96 (death counted as ≥400). Secondary efficacy outcomes were VL <60 and <1000 copies/ml at week-96, death/WHO 3/4 events, changes in CD4 (count/percentage), and genotypic resistance (assays ongoing); safety outcomes were grade 3/4, serious, and ART-modifying adverse events (AEs); changes in total, low density lipoprotein (LDL) and high-density lipoprotein (HDL) cholesterol and triglycerides; and changes in bilirubin and creatinine clearance (CrCl). Other outcomes included changes in weight-, height- and body mass index (BMI)-for-age and bone mineral density Z-scores. In a within-trial cost-effectiveness analysis over the 96 weeks of the trial, health was estimated using quality-adjusted life-years (QALYs) and costs were estimated from the health-system perspective and included ART costs, clinic visits and hospital stays in 2022 US dollars, both discounted at 3% per annum.

The sample size of 920 children (including 2.5% loss to follow-up; reduced from 10% in the original protocol) provided 88% power to demonstrate ATV/r was non-inferior (12% margin) to LPV/r (two-sided alpha=5%), assuming 80% VL <400 copies/ml at week-96, and 89% power to detect 10% higher VL <400 copies/ml in each of DTG and DRV/r than LPV/r and ATV/r combined (two-sided alpha=3%; as multiple comparisons). An independent data monitoring committee reviewed the interim data at four meetings using the Haybittle–Peto criterion (99.9% confidence intervals).

Analyses were intention-to-treat. Analyses of the primary endpoint (VL <400 copies/ml), used logistic regression (adjusting for stratification factors), then marginal estimation of risk differences. For non-inferiority comparisons, secondary per-protocol analyses included children who received randomised anchor drug for >90% of follow-up. Sub-group analysis used interaction tests. For VL <60 and <1000 copies/ml, analysis was similar. For death/WHO 3/4 events, and grade 3/4, serious and ART-modifying AEs, groups were compared using Cox regression (unadjusted). Changes in continuous outcomes were analysed using Normal generalized estimating equations adjusting for visit, and stratification factors and baseline (and interactions between these factors and visit), for an overall test of difference between groups over all visits (independent correlation). Analysis was conducted using Stata (version 17.0). The 95% confidence intervals were not adjusted for multiple testing.

The funder, European Developing Country Clinical Trial Partnership (EDCTP), and pharmaceutical companies donating additional funding (Gilead Sciences, Johnson and Johnson) and trial medications (ViiV Healthcare, Gilead sciences, Johnson and Johnson, CIPLA) did not participate in design, conduct or analysis of the trial.

## Results

919 children were randomised between 17 December 2018 and 1 April 2021 and followed for minimum 96 weeks. 227 were randomised to LPV/r, 231 ATV/r, 232 DRV/r and 229 to DTG (Figure 1). Baseline characteristics were similar between arms (Table 1; Table S1 in Supplementary Appendix 1). 497(54%) children were male; median age was 10 years (IQR 8,13); 777 (84.5%) had WHO stage 1/2 disease. Median weight-height- and BMI-for-age Z-scores were between -1 and -1.6. Median VL was 17,573 copies/mL (IQR 5549,55700); CD4 count 669 cells/mm^3^ (413,971) and CD4% 28% (19%,36%). Median time on first-line ART was 5.6 years (44% nevirapine, 56% efavirenz).

**Figure 1.**
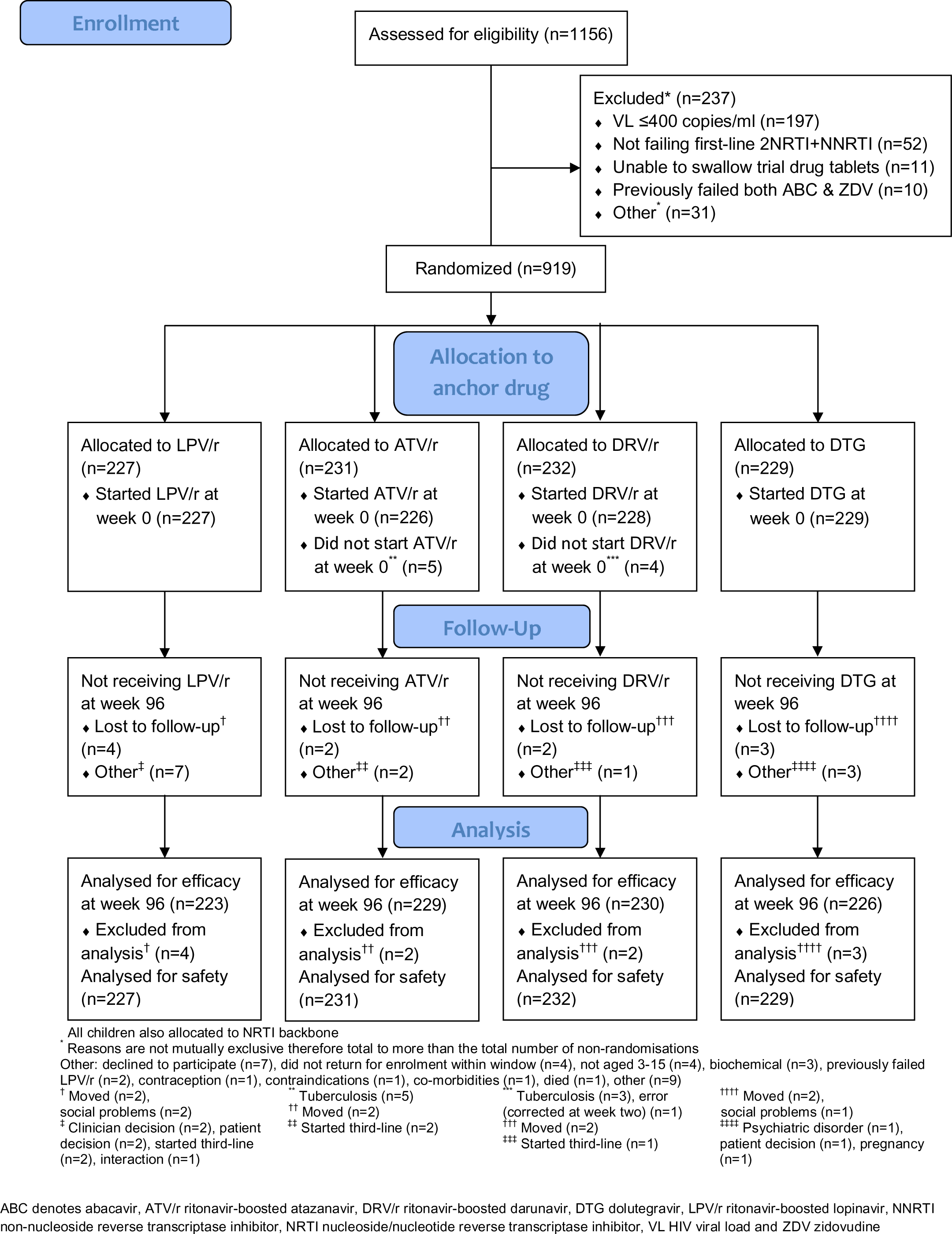
CONSORT flow diagram

**Table 1:**
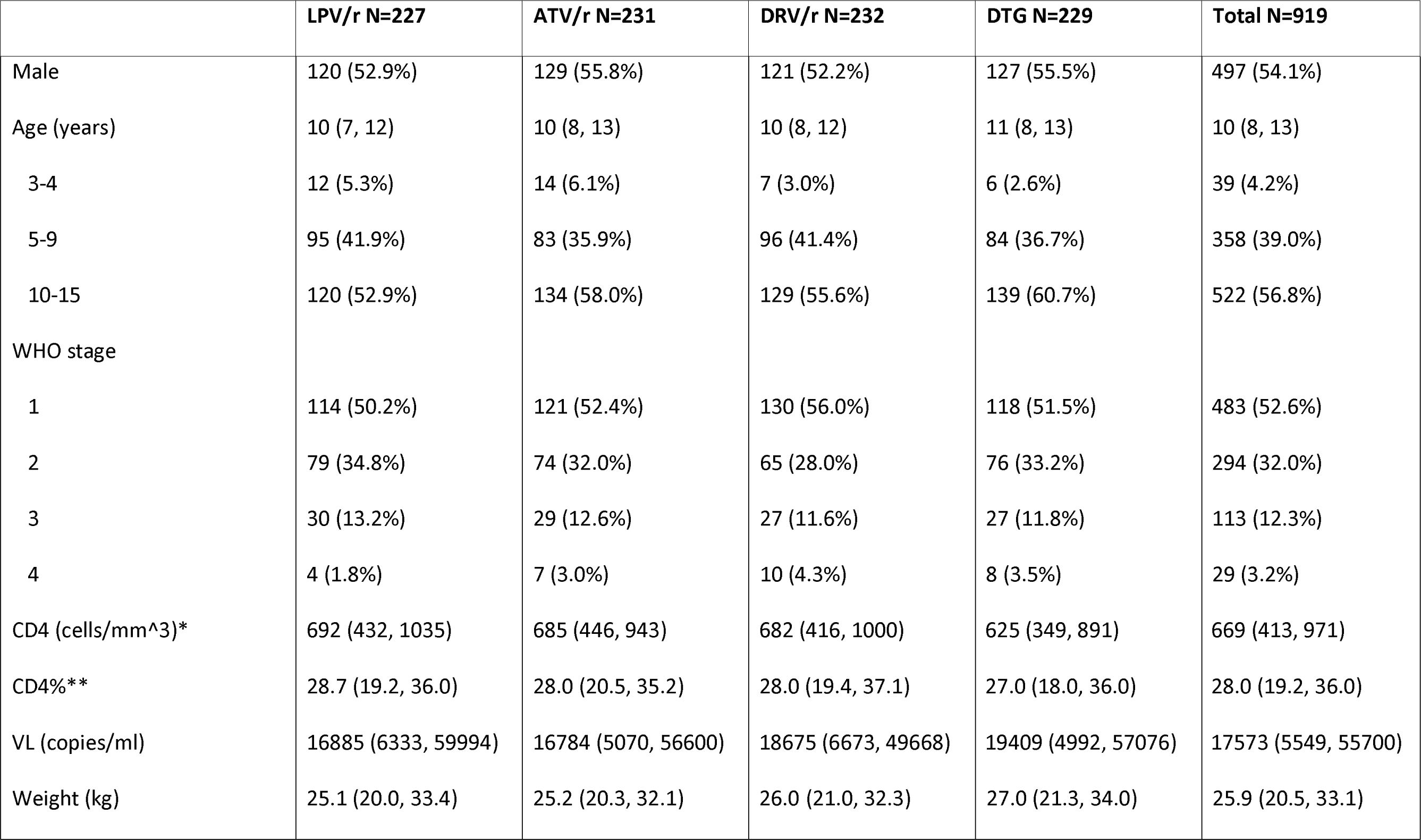
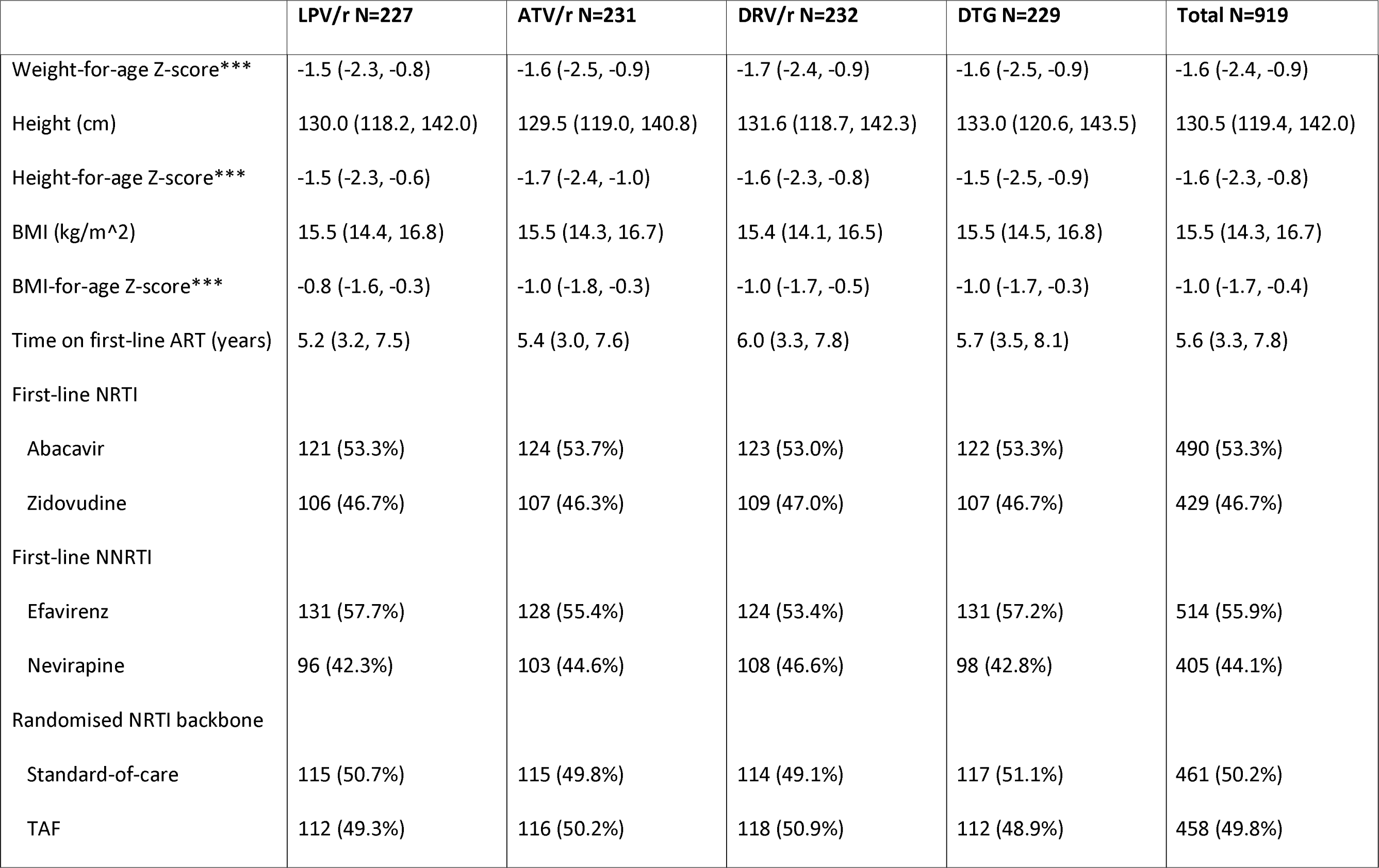
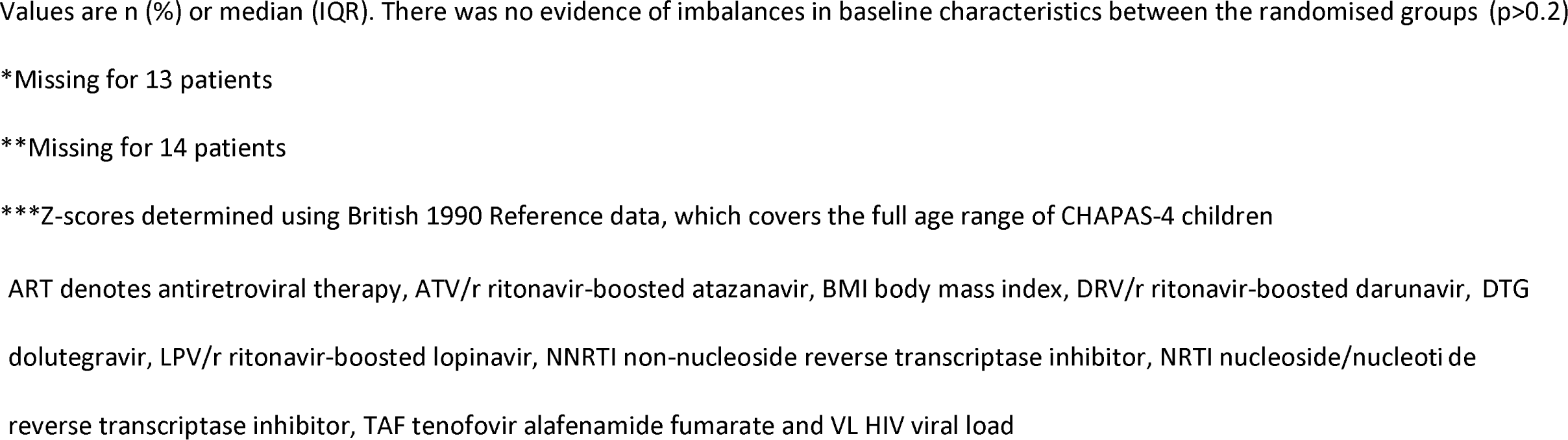
Baseline characteristics.

|  | **LPV/r N=227** | **ATV/r N=231** | **DRV/r N=232** | **DTG N=229** | **Total N=919** |
| --- | --- | --- | --- | --- | --- |
| Male | 120 (52.9%) | 129 (55.8%) | 121 (52.2%) | 127 (55.5%) | 497 (54.1%) |
| Age (years) | 10 (7, 12) | 10 (8, 13) | 10 (8, 12) | 11 (8, 13) | 10 (8, 13) |
| 3-4 | 12 (5.3%) | 14 (6.1%) | 7 (3.0%) | 6 (2.6%) | 39 (4.2%) |
| 5-9 | 95 (41.9%) | 83 (35.9%) | 96 (41.4%) | 84 (36.7%) | 358 (39.0%) |
| 10-15 | 120 (52.9%) | 134 (58.0%) | 129 (55.6%) | 139 (60.7%) | 522 (56.8%) |
| WHO stage |  |  |  |  |  |
| 1 | 114 (50.2%) | 121 (52.4%) | 130 (56.0%) | 118 (51.5%) | 483 (52.6%) |
| 2 | 79 (34.8%) | 74 (32.0%) | 65 (28.0%) | 76 (33.2%) | 294 (32.0%) |
| 3 | 30 (13.2%) | 29 (12.6%) | 27 (11.6%) | 27 (11.8%) | 113 (12.3%) |
| 4 | 4 (1.8%) | 7 (3.0%) | 10 (4.3%) | 8 (3.5%) | 29 (3.2%) |
| CD4 (cells/mm<sup>3</sup>)* | 692 (432, 1035) | 685 (446, 943) | 682 (416, 1000) | 625 (349, 891) | 669 (413, 971) |
| CD4%** | 28.7 (19.2, 36.0) | 28.0 (20.5, 35.2) | 28.0 (19.4, 37.1) | 27.0 (18.0, 36.0) | 28.0 (19.2, 36.0) |
| VL (copies/ml) | 16885 (6333, 59994) | 16784 (5070, 56600) | 18675 (6673, 49668) | 19409 (4992, 57076) | 17573 (5549, 55700) |
| Weight (kg) | 25.1 (20.0, 33.4) | 25.2 (20.3, 32.1) | 26.0 (21.0, 32.3) | 27.0 (21.3, 34.0) | 25.9 (20.5, 33.1) |
| Weight-for-age Z-score*** | -1.5 (-2.3, -0.8) | -1.6 (-2.5, -0.9) | -1.7 (-2.4, -0.9) | -1.6 (-2.5, -0.9) | -1.6 (-2.4, -0.9) |
| Height (cm) | 130.0 (118.2, 142.0) | 129.5 (119.0, 140.8) | 131.6 (118.7, 142.3) | 133.0 (120.6, 143.5) | 130.5 (119.4, 142.0) |
| Height-for-age Z-score*** | -1.5 (-2.3, -0.6) | -1.7 (-2.4, -1.0) | -1.6 (-2.3, -0.8) | -1.5 (-2.5, -0.9) | -1.6 (-2.3, -0.8) |
| BMI (kg/m^2) | 15.5 (14.4, 16.8) | 15.5 (14.3, 16.7) | 15.4 (14.1, 16.5) | 15.5 (14.5, 16.8) | 15.5 (14.3, 16.7) |
| BMI-for-age Z-score*** | -0.8 (-1.6, -0.3) | -1.0 (-1.8, -0.3) | -1.0 (-1.7, -0.5) | -1.0 (-1.7, -0.3) | -1.0 (-1.7, -0.4) |
| Time on first-line ART (years) | 5.2 (3.2, 7.5) | 5.4 (3.0, 7.6) | 6.0 (3.3, 7.8) | 5.7 (3.5, 8.1) | 5.6 (3.3, 7.8) |
| First-line NRTI |  |  |  |  |  |
| Abacavir | 121 (53.3%) | 124 (53.7%) | 123 (53.0%) | 122 (53.3%) | 490 (53.3%) |
| Zidovudine | 106 (46.7%) | 107 (46.3%) | 109 (47.0%) | 107 (46.7%) | 429 (46.7%) |
| First-line NNRTI |  |  |  |  |  |
| Efavirenz | 131 (57.7%) | 128 (55.4%) | 124 (53.4%) | 131 (57.2%) | 514 (55.9%) |
| Nevirapine | 96 (42.3%) | 103 (44.6%) | 108 (46.6%) | 98 (42.8%) | 405 (44.1%) |
| Randomised NRTI backbone |  |  |  |  |  |
| Standard-of-care | 115 (50.7%) | 115 (49.8%) | 114 (49.1%) | 117 (51.1%) | 461 (50.2%) |
| TAF | 112 (49.3%) | 116 (50.2%) | 118 (50.9%) | 112 (48.9%) | 458 (49.8%) |

Values are n (%) or median (IQR). There was no evidence of imbalances in baseline characteristics between the randomised groups ( $p>0.2$ )
\*Missing for 13 patients
\*\*Missing for 14 patients
\*\*\*Z-scores determined using British 1990 Reference data, which covers the full age range of CHAPAS-4 children
ART denotes antiretroviral therapy, ATV/r ritonavir-boosted atazanavir, BMI body mass index, DRV/r ritonavir-boosted darunavir, DTG dolutegravir, LPV/r ritonavir-boosted lopinavir, NNRTI non-nucleoside reverse transcriptase inhibitor, NRTI nucleoside/nucleotide reverse transcriptase inhibitor, TAF tenofovir alafenamide fumarate and VL HIV viral load

At randomisation, 910/919 (99.0%) children initiated their randomised anchor drug (eight randomised to ATV/r or DRV/r initiated LPV/r or DTG because of tuberculosis (protocol-specified modification), one error). Over 96 weeks, 98.9% of visits were attended and only 11 (1.2%) children were lost to follow-up. 674 children (73.3%) entered extended follow-up (median 60 (IQR 30,75) additional weeks). Through week 96, children spent 98.6% of follow-up on allocated anchor drug (99.1% DTG, 98.5% DRV/r, 98.6% ATV/r, 98.4% LPV/r) and only five (0.5%) children initiated third-line ART (2 LPV/r, 2 ATV/r, 1 DRV/r, 0 DTG). In extended follow-up, children spent 86.2% of time on allocated anchor drug (99.1% DTG, 95.6% DRV/r, 93.7% ATV/r, 54.9% LPV/r) (Figure S1 in Supplementary Appendix 1).

At week-96, 92.0% DTG, 88.3% DRV/r, 84.3% ATV/r and 80.7% LPV/r had VL <400 copies/mL (Figure 2). Considering the pre-specified comparisons (Table S2 in Supplementary Appendix 1), DTG was superior to LPV/r and ATV/r arms combined (adjusted difference 9.7% [95% confidence interval (CI) 4.8,14.5]; p<0.0001). DRV/r was non-inferior to LPV/r and ATV/r arms combined and showed a trend to superiority (adjusted difference 5.6% [0.3,11.0]; p=0.04); vs. threshold p=0.03 due to multiple comparisons). ATV/r was non-inferior to LPV/r (adjusted difference +3.4% [-3.4,10.2]; p=0.33). In a post-hoc analysis, DTG showed a trend to superiority vs. DRV/r (adjusted difference 4.0% [-1.3,9.4]; p=0.14). Per-protocol analyses of non-inferiority comparisons were similar: 88.4% DRV/r vs. 82.7% LPV/r and ATV/r combined had VL <400 copies/mL (adjusted difference 5.4% [-0.0,10.7]; p=0.051); 83.9% ATV/r vs. 81.4% LPV/r had VL <400copies/mL (adjusted difference 2.1% [-4.8,8.9]; p=0.55). There was no evidence of heterogenicity among 11 prespecified sub-groups (p**_interaction_**>0.05), including first-line NRTI (ABC vs. ZDV), randomised NRTI backbone, country and baseline VL, apart from a minimal difference in VL response for DTG vs. LPV/r and ATV/r combined based on first-line nevirapine or efavirenz use (Figures S2-S4 in Supplementary Appendix 1). Results using other viral load thresholds (<60 and <1000 copies/ml) were similar, as was suppression at weeks 48 and 144 (Figure 2; Table S2 in Supplementary Appendix 1).

**Figure 2.**
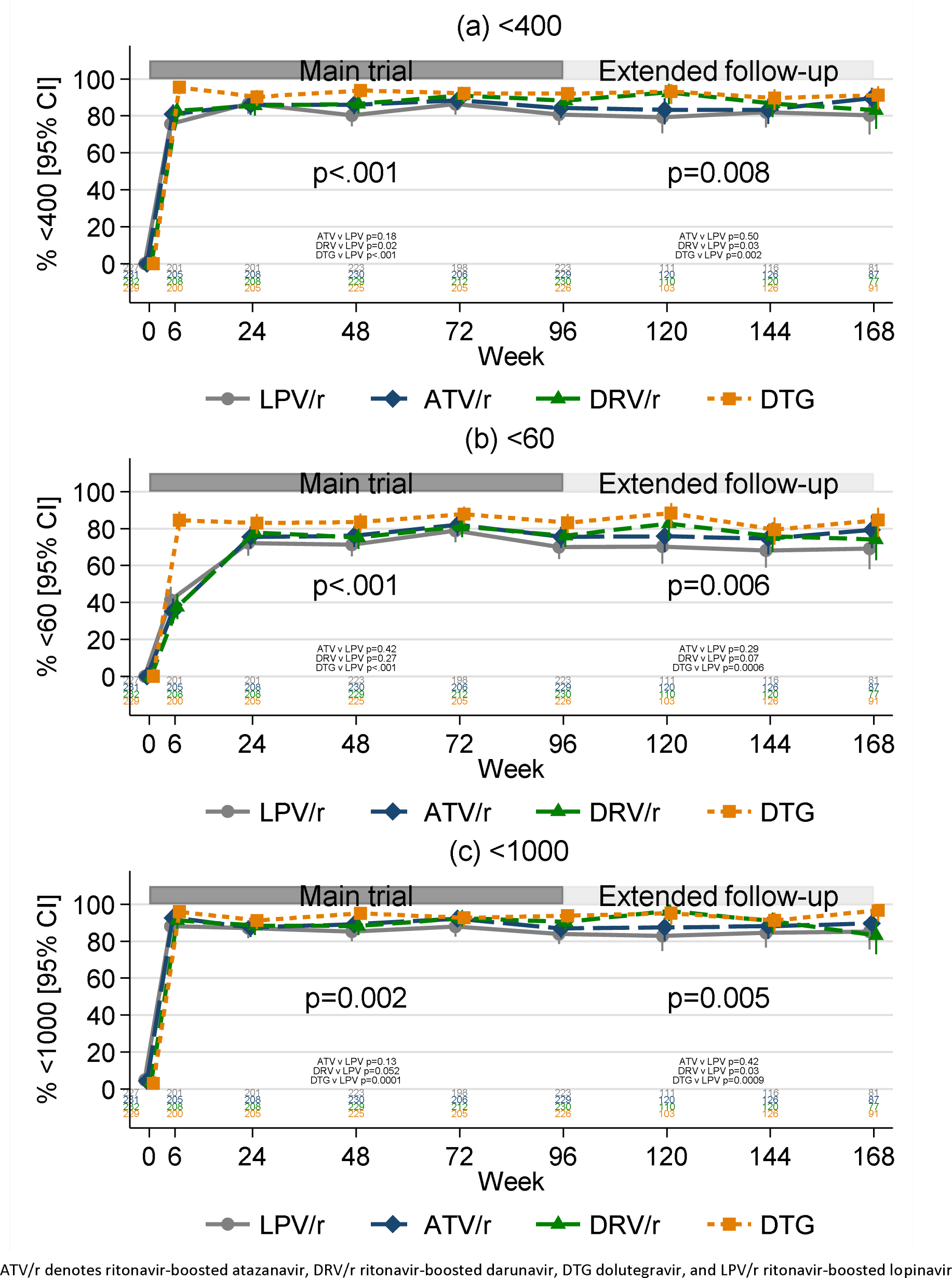
Percentage of children with HIV viral load (copies/ml) <400 copies/ml (a), <60 copies/ml (b) and <1000 copies/ml (c), over time during the main trial and during extended follow-up

Over 96 weeks, there were only nine WHO stage 3/4 events (5 DTG, 2 DRV/r, 1 ATV/r, 2 LPV/r) and one death in the DTG arm from hypotension/toxic shock secondary to severe malnutrition, judged unrelated to ART (Table S3 Supplementary Appendix 1).

CD4 count improved in all arms (Figure S5 in Supplementary Appendix 1) with no evidence of differences between arms over 96-weeks and in extended follow-up.

Weight- and BMI-for-age Z-scores increased significantly more with ATV/r, DRV/r and DTG vs. LPV/r (Figure 3). There was no evidence that anchor drugs effects on weight-for-age-Z-scores differed according to backbone randomisation (p**_interactio_**=**_n_** 0.51) (Figure S6, Table S6 in Supplementary Appendix 1).

**Figure 3.**
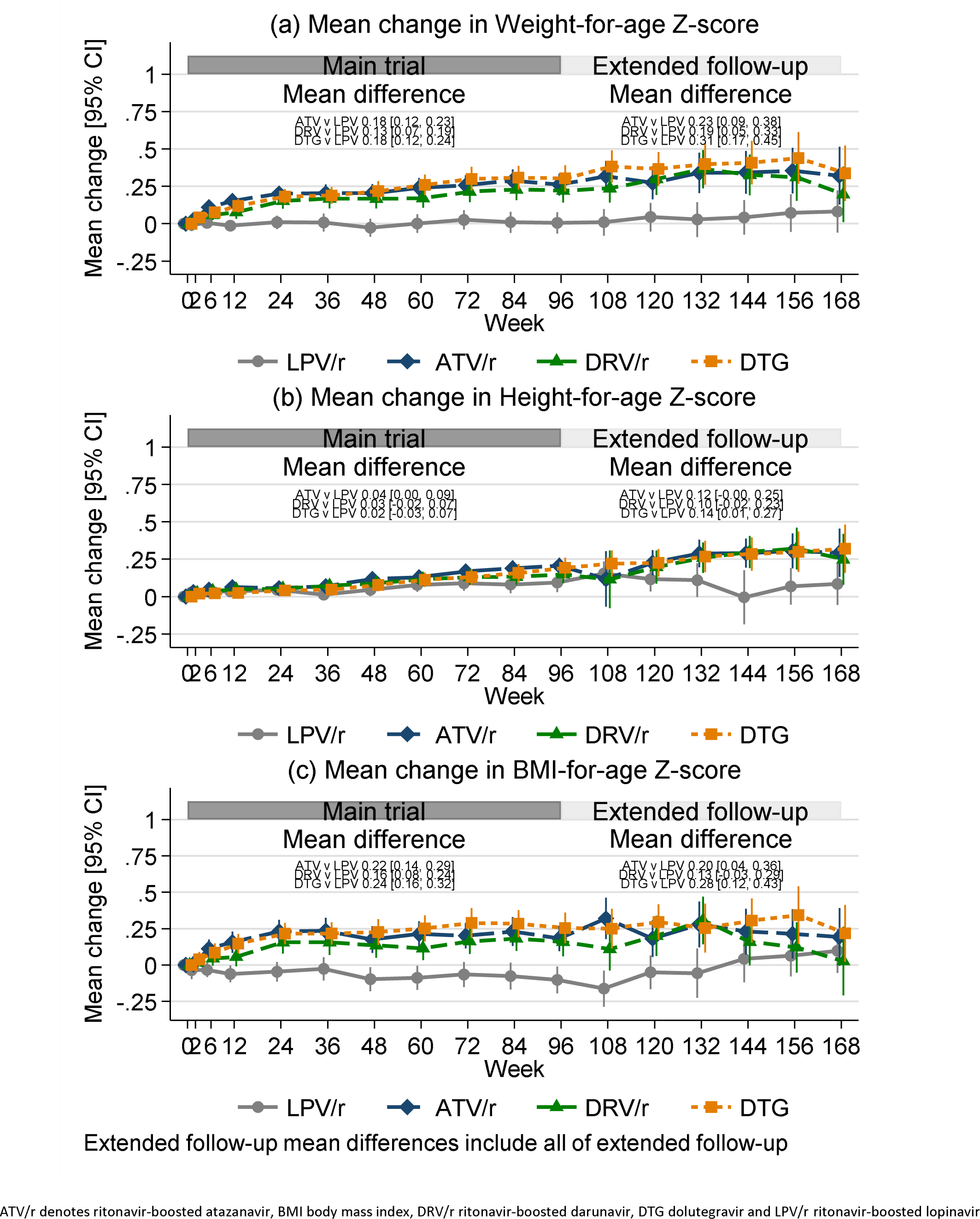
Change in (a) weight-, (b) height- and (c) BMI-for-age Z-scores

Over 96 weeks, 127/919 (13.8%) children experienced a total of 176 grade 3/4 adverse events (AEs) (Table 2; Table S4 in Supplementary Appendix 1), most commonly hyperbilirubinemia almost exclusively with ATV/r, and consistent with expected effects on bilirubin (Figure S7 in Supplementary Appendix 1). Fewer children experienced grade 3/4 AEs with DTG (5.2%) vs. LPV/r (11.5%) (p=0.02); there was no evidence of differences between DRV/r (8.6%) vs. LPV/r (11.5%) (p=0.31).

**Table 2:**
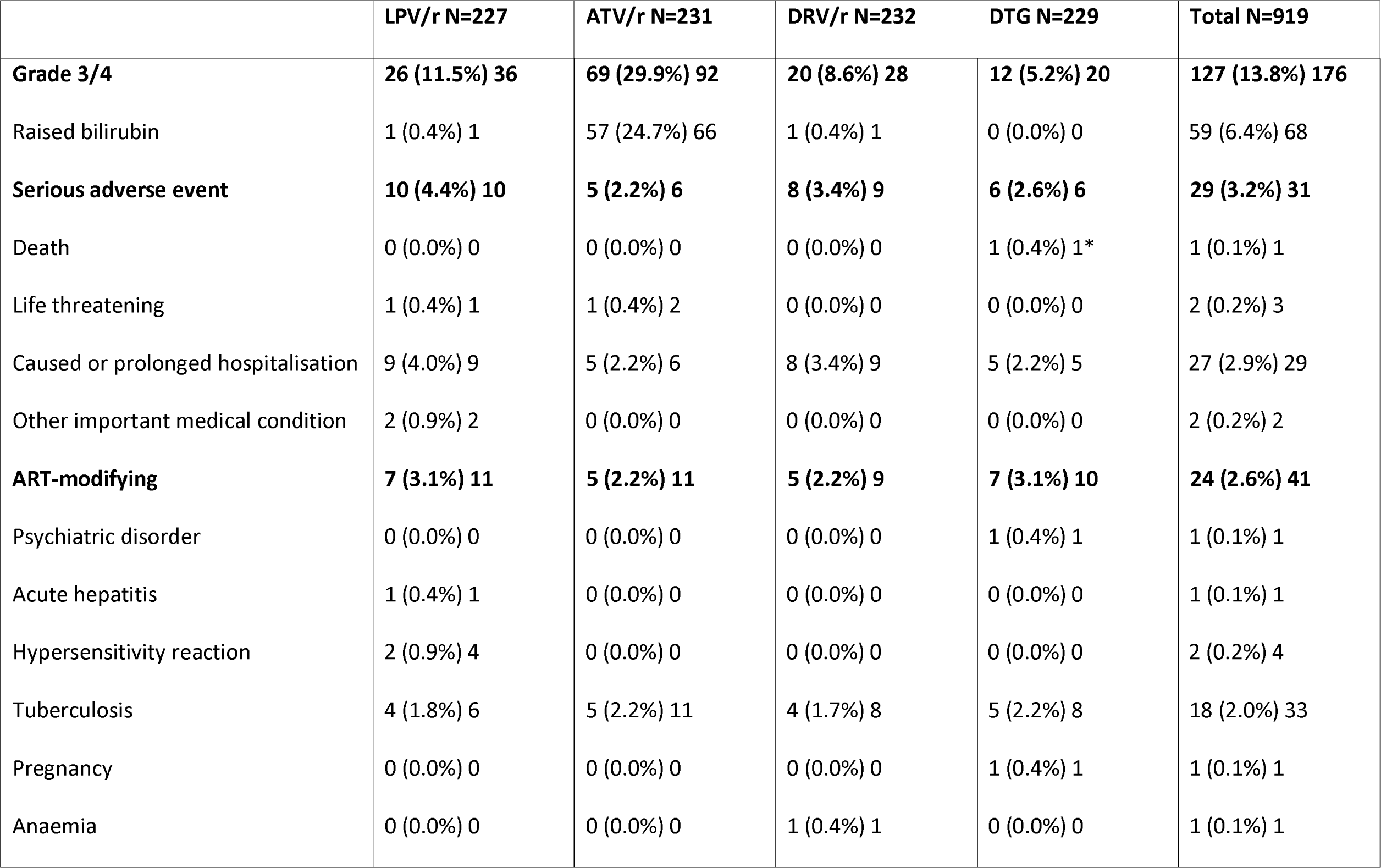
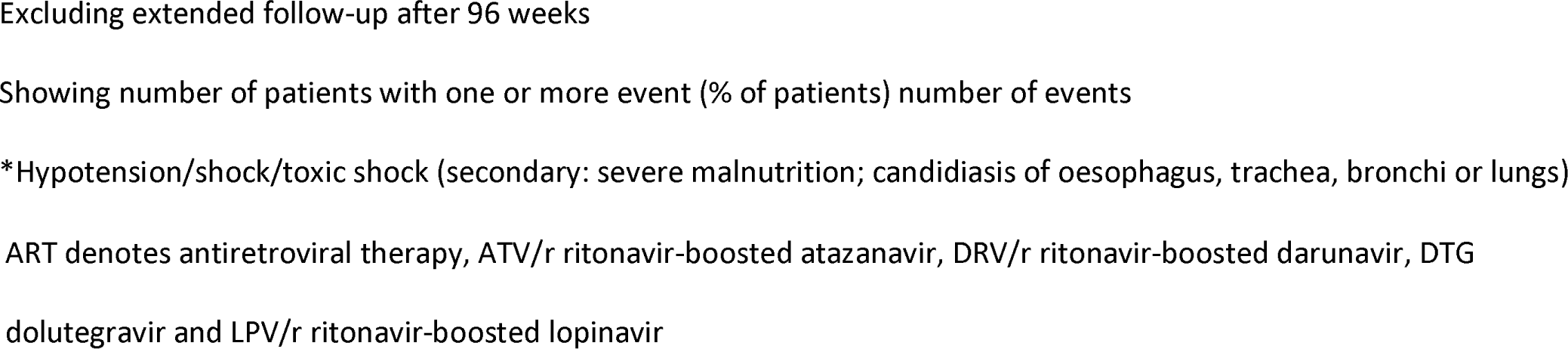
Grade 3 and 4, serious and ART-modifying adverse events during 96-week follow-up.

|  | LPV/r N=227 | ATV/r N=231 | DRV/r N=232 | DTG N=229 | Total N=919 |
| --- | --- | --- | --- | --- | --- |
| **Grade 3/4** | **26 (11.5%) 36** | **69 (29.9%) 92** | **20 (8.6%) 28** | **12 (5.2%) 20** | **127 (13.8%) 176** |
| Raised bilirubin | 1 (0.4%) 1 | 57 (24.7%) 66 | 1 (0.4%) 1 | 0 (0.0%) 0 | 59 (6.4%) 68 |
| **Serious adverse event** | **10 (4.4%) 10** | **5 (2.2%) 6** | **8 (3.4%) 9** | **6 (2.6%) 6** | **29 (3.2%) 31** |
| Death | 0 (0.0%) 0 | 0 (0.0%) 0 | 0 (0.0%) 0 | 1 (0.4%) 1* | 1 (0.1%) 1 |
| Life threatening | 1 (0.4%) 1 | 1 (0.4%) 2 | 0 (0.0%) 0 | 0 (0.0%) 0 | 2 (0.2%) 3 |
| Caused or prolonged hospitalisation | 9 (4.0%) 9 | 5 (2.2%) 6 | 8 (3.4%) 9 | 5 (2.2%) 5 | 27 (2.9%) 29 |
| Other important medical condition | 2 (0.9%) 2 | 0 (0.0%) 0 | 0 (0.0%) 0 | 0 (0.0%) 0 | 2 (0.2%) 2 |
| **ART-modifying** | **7 (3.1%) 11** | **5 (2.2%) 11** | **5 (2.2%) 9** | **7 (3.1%) 10** | **24 (2.6%) 41** |
| Psychiatric disorder | 0 (0.0%) 0 | 0 (0.0%) 0 | 0 (0.0%) 0 | 1 (0.4%) 1 | 1 (0.1%) 1 |
| Acute hepatitis | 1 (0.4%) 1 | 0 (0.0%) 0 | 0 (0.0%) 0 | 0 (0.0%) 0 | 1 (0.1%) 1 |
| Hypersensitivity reaction | 2 (0.9%) 4 | 0 (0.0%) 0 | 0 (0.0%) 0 | 0 (0.0%) 0 | 2 (0.2%) 4 |
| Tuberculosis | 4 (1.8%) 6 | 5 (2.2%) 11 | 4 (1.7%) 8 | 5 (2.2%) 8 | 18 (2.0%) 33 |
| Pregnancy | 0 (0.0%) 0 | 0 (0.0%) 0 | 0 (0.0%) 0 | 1 (0.4%) 1 | 1 (0.1%) 1 |
| Anaemia | 0 (0.0%) 0 | 0 (0.0%) 0 | 1 (0.4%) 1 | 0 (0.0%) 0 | 1 (0.1%) 1 |

Excluding extended follow-up after 96 weeks
Showing number of patients with one or more event (% of patients) number of events
\*Hypotension/shock/toxic shock (secondary: severe malnutrition; candidiasis of oesophagus, trachea, bronchi or lungs)
ART denotes antiretroviral therapy, ATV/r ritonavir-boosted atazanavir, DRV/r ritonavir-boosted darunavir, DTG dolutegravir and LPV/r ritonavir-boosted lopinavir

Twenty-nine (3.2%) children experienced 31 serious adverse events (SAEs) (6 DTG, 8 DRV/r, 5 ATV/r, 10 LPV/r) (p>0.1) (Table S5 in Supplementary Appendix 1); most were hospitalisations, with intercurrent infections. Twenty-four (2.6%) children experienced a total of 41 ART-modifying AEs of any grade, with no evidence of difference across arms (7 DTG, 5 DRV/r, 5 ATV/r, 7 LPV/r) (p>0.5), most commonly (33) for protocol-specified modifications due to tuberculosis.

Over 96 weeks, creatinine clearance decreased more with DTG vs. LPV/r, ATV/r and DRV/r (p<0.001), although differences were small (in the order of 2ml/min) and within the normal range (Figure S8 in Supplementary Appendix 1). Total, HDL and LDL cholesterol increased significantly more with LPV/r vs. ATV/r, DRV/r or DTG (all p<0.001; Figure S9 in Supplementary Appendix 1). Triglycerides increased in all arms up to 48 weeks, more markedly in the LPV/r arm (overall p=0.002), then decreased through to week-96.

Dual-energy X-ray absorptiometry (DEXA) scans were undertaken at weeks 0, 48 and 96 in a subset of 170 children. At baseline, low (Z-score ≤2) total body-less-head (TBLH) BMD was observed in 28 (18%) children, 21(13%) had low lumbar spine (LS) total BMD, and 15 (9%) had both.^7^ At week-96, TBLH bone mineral content (BMC) and BMD increased more with ATV/r, DRV/r and DTG vs. LPV/r (p=0.02 and p=0.0003, respectively) (Figure S10 in Supplementary Appendix 1). TBLH BMD Z-scores also decreased most on LPV/r vs. the other three arms (p=0.001); lumbar total BMD Z-score (p=0.01) decreased slightly less with DRV/r vs. LPV/r, ATV/r and DTG. There was no evidence of differences in lumbar total BMC and BMD (p>0.2). Of note the majority of children on SOC remained on ABC or ZDV throughout and only 7 (1.5%) children switched to TDF/TAF over 96 weeks.

There was no evidence of difference in QALYs across the four anchor drug arms, so the main health economics analysis focused on cost differences. DTG was the least costly, saving $190.77 compared to ATZ/r, which was the second least costly, while DRV/r was the most expensive. The probability of DTG being least costly was 100%. Further detail is included on cost effectiveness in Supplementary Appendix 2.

## Discussion

CHAPAS-4 is the largest clinical trial to evaluate second-line ART treatment in children in Africa. DTG-based regimens were superior with respect to virologic suppression compared with LPV/r and ATV/r arms combined; DRV/r also showed a trend towards superior virological efficacy compared to LPV/r and ATV/r arms combined, and in a posthoc analysis, DTG showed a trend to superiority to DRV/r. These comparisons between the four main currently available second-line anchor drugs for children provide much-needed robust evidence to guide future drug formulation development and paediatric ART guidelines. LPV/r was associated with the poorest virological outcomes and with poorer growth, lipid profiles and bone health.

The superior virologic suppression with DTG vs. bPIs confirms and extends findings from the ODYSSEY trial which showed superiority of DTG vs. SOC for both first- and second-line ART (ODYSSEY second-line SOC being 72% LPV/r, 24% ATV/r, 1% DRV/r).^8^ CHAPAS-4 provides additional evidence through direct randomised comparisons of DTG and DRV/r vs. ATV/r or LPV/r. Given the cost-effectiveness of DTG, its small milligram dosing and authorisation for use below 3 years, these results further support DTG as anchor drug of choice in second-line regimens in World Health Organisation (WHO) guidelines.^9^ WHO recommended DTG in combination with an optimised NRTI backbone for adults experiencing NNRTI-based first-line ART failure,^9^ partly based on the DAWNING trial which demonstrated superior efficacy and safety of DTG vs. LPV/r in combination with two NRTIs.^10^ Further evidence for this recommendation came from the NADIA study which showed that DTG was non-inferior to boosted DRV in combination with TDF or ZDV (90% vs. 87% viral suppression at 96 weeks).^11^

CHAPAS-4 demonstrated immune reconstitution for all four anchor drugs, particularly in the first 24 weeks after switching to second-line ART (Figure S5 in Supplementary Appendix 1). Age-appropriate weight gains were observed with all drugs except LPV/r, which showed minimal increases in weight-for-age Z-scores in a population with already low z-scores at baseline (Figure 3). A systematic review and meta-analysis evaluating weight gain among adults reported greater weight gain among those receiving DTG with TAF compared to other NRTIs (ABC/ZDV with 3TC/emtricitabine)^12^ but we did not observe excessive weight gain associated with any anchor drug/NRTI combination, including DTG/TAF. The excess weight gain in adults has been associated with advanced immunosuppression at ART initiation, high HIV viral load, female sex and black race, mostly occurring in the first 2 years of therapy.^13^ This phenomena has been described as a “return to health” where resting energy expenditure returns to normal as HIV viremia and inflammation are contolled.^14^ Participants in CHAPAS-4 were either normal or underweight (Table 1), and none had evidence of obesity at randomisation, so findings from CHAPAS-4 may not generalise to more obese populations.

All anchor drugs were well tolerated, with most participants remaining on their allocated anchor drug through week-96. As expected, lipid profiles were less favourable for children on LPV/r vs. other anchor drugs and raised bilirubin was associated with ATV/r.

These findings also show that DRV/r and ATV/r are safe, effective once-daily treatment options which can be considered if DTG cannot be used. Previous small studies have shown ATV/r to be both safe and effective in children and potentially a preferred and better tolerated second-line option compared to LPV/r,^15^ as long as hyperbilirubinaemia either does not occur or is acceptable. LPV/r use in children has had considerable challenges of palatability and twice-daily dosing. The additional data on poorer growth, higher toxicity and poorer virological outcomes in CHAPAS-4 emphasize that LPV/r is now a suboptimal option.

One strength of CHAPAS-4 is its power to compare both DTG and DRV/r with ATV/r and LPV/r while employing a factorial design to further compare the impact of NRTI backbone. Although power to test interactions is more limited, there was no evidence of abnormal weight gain with any combination of NRTIs and anchor drugs. Loss-to-follow-up was minimal, despite the COVID-19 pandemic. Whilst the findings can be generalised to current policy in children and adolescents requiring second-line therapy after NNRTI-based first-line ART, children currently initiating first-line DTG may also require robust second-line options. One limitation is that CHAPAS-4 does not provide direct evidence for second-line bPIs in this situation; however, safety and efficacy of DRV/r or ATV/r could be inferred and they will remain important options in future. One factor that may have impacted efficacy of the PI options requiring ritonavir boosting was the lack of co-formulated tablets, resulting in a relatively high pill burden (although a small 25mg ritonavir generic pill was used). Overcoming this barrier may further enhance the effectiveness of bPIs in children in the future.

Overall, CHAPAS-4 results confirm the superior and sustained efficacy, toxicity and cost-effectiveness of DTG compared with bPIs. Boosted DRV and ATV were also efficacious and well-tolerated anchor drug options. This provides essential evidence that will inform global policy and guidelines in prioritising the development of cost-effective paediatric formulations for roll out in Africa and globally.

## Data Availability

MRC CTU at UCL supports a controlled access approach based on completion of a data request proforma available from the corresponding author (mrcctu.ucl.ac.uk/our-research/other-research-policy/data-sharing).

## Acknowledgements

We thank the participants and their families for taking part in the trial. We also acknowledge the following individuals in the partner institutions, funding bodies and pharmaceutical companies.

Clinical Trials Unit:

MRC CTU at UCL

Di Gibb, Sarah Walker, Anna Turkova, Clare Shakeshaft, Moira Spyer, Margaret Thomason, Anna Griffiths, Lara Monkiewicz, Sue Massingham, Alex Szubert, Alasdair Bamford, Katja Doerholt, Amanda Bigault, Nimisha Dudakia, Annabelle South, Nadine Van Looy, Carly Au, Hannah Sweeney

Trial Sites:

Joint Clinical Research Centre Lubowa, Uganda: Cissy M. Kityo, Victor Musiime, Eva Natukunda, Esether Nambi, Diana Rutebarika Antonia, Rashida Nazzinda, Imelda Namyalo, Joan Nangiya, Lilian Nabeeta, Aidah Nakalyango, Lilian Kobusingye, Caroline Otike, Winnie Namala, Phionah Ampaire, Ayesiga Edgar, Claire Nasaazi, Milly Ndigendawani, Paul Ociti, Priscilla Kyobutungi, Ritah Mbabazi, Phyllis Mwesigwa Rubondo, Juliet Ankunda, Mariam Naabalamba, Mary Nannungi, Alex Musiime, Faith Mbasani, Babu Enoch Louis, Josephine Namusanje, Denis Odoch, Edward Bagirigomwa, Eddie Rubanga, Disan Mulima, Paul Oronon, Eram David Williams, David Baliruno, Josephine Kobusingye, Agnes Uyungrwoth, Barbara Mukanza, Jimmy Okello, Emily Ninsiima, Lutaro Ezra, Christine Nambi, Nansaigi Mangadalen, Musumba Sharif, Nobert B. Serunjogi, Otim Thomas

Joint Clinical Research Centre Mbarara, Uganda: Abbas Lugemwa, Shafic Makumbi, Sharif Musumba, Edward Mawejje, Ibrahim Yawe, Linda Jovia Kyomuhendo, Mariam Kasozi, Rogers Ankunda, Samson kariisa, Christine Inyakuwa, Emily Ninsiima, Lorna Atwine, Beatrice Tumusiime, John Ahuura, Deogracious Tukwasibwe, Violet Nagasha, Judith Kukundakwe, Mariam Zahara Nakisekka, Ritah Winnie Nambejja, Mercy Tukamushaba, Rubinga Baker, Edridah Keminyeto, Barbara Ainebyoona, Sula Myalo, Juliet Acen, Nicholas Jinta Wangwe, Ian Natuhurira, Gershom Kananura Natukunatsa University Teaching Hospital, Zambia: Veronica Mulenga, Chishala Chabala, Joyce Chipili Lungu, Monica Kapasa, Khozya Zyambo, Kevin Zimba, Chungu Chalilwe, Dorothy Zangata, Ellen Shingalili, Naomi Mumba, Nayunda Kaonga, Mukumbi Kabesha, Oliver Mwenechanya, Terrence Chipoya, Friday Manakalanga, Stephen Malama, Daniel Chola

Arthur Davison Children’s Hospital, Zambia: Bwendo Nduna, Mwate Mwamabazi, Kabwe Banda, Beatrice Kabamba, Muleya Inambao, Pauline Mahy Mukandila, Mwizukanji Nachamba, Stella Himabala, Shadrick Ngosa, Davies Sondashi, Collins Banda, Mark Munyangabe, Grace Mbewe Ngoma, Sarah Chimfwembe, Mercy Lukonde Malasha, Mumba Kajimalwendo, Henry Musukwa, Shadrick Mumba

University of Zimbabwe Clinical Research Centre, Zimbabwe: James Hakim, Mutsa Bwakura-Dangarembizi, Kusum Nathoo, Taneal Kamuzungu, Ennie Chidziva, Joyline Bhiri, Joshua Choga, Hilda Angela Mujuru, Godfrey Musoro, Vivian Mumbiro, Moses Chitsamatanga, Constantine Mutata, Shepherd Mudzingwa, Secrecy Gondo, Columbus Moyo, Ruth Nhema, Kathryn Boyd, Farai Matimba, Vinie Kouamou, Richard Matarise, Zorodzai Tangwena, Taona Mudzviti, Allen Matubu, Alfred Kateta, Victor Chinembiri, Dorinda Mukura, Joy Chimanzi, Dorothy Murungu, Wendy Mapfumo, Pia Ngwaru, Lynette Chivere, Prosper Dube, Trust Mukanganiki, Sibusisiwe Weza, Tsitsi Gwenzi, Shirley Mutsai, Misheck Phiri, Makhosonke Ndlovu, Tapiwa Gwaze, Stuart Chitongo, Winisayi Njaravani, Sandra Musarurwa, Cleopatra Langa, Sue Tafeni, Wilbert Ishemunyoro, Nathalie Mudzimirema Mpilo Central Hospital, Zimbabwe: Wedu Ndebele, Mary Nyathi, Grace Siziba, Getrude Tawodzera, Tracey Makuchete, Takudzwa Chidarura, Shingaidzo Murangandi, Lawrence Mafaro, Owen Chivima, Sifiso Dumani, Beaullar Mampondo, Constance Maphosa, Debra Mwale, Rangarirai Dhlamini, Thabani Sibanda, Nobukhosi Madubeko, Silibaziso Nyathi, Zibusiso Matiwaza, Blessing Sanyanga, Prince Ziyera, Gamuchirai Mauro, Titshabona Ncube, Again Gwapedza, Davison Mashoko Local External Site Monitors

Uganda: Sylvia Nabukenya, Harriet Tibakabikoba, Sarah Nakalanzi, Cynthia Williams

Zimbabwe: Precious Chandiwana, Winnie Gozhora, Benedictor Dube

Zambia: Sylvia Mulambo, Hope Mwanyungwi

Sub-studies

PK sub-studies – Radboud University Medical Centre: David Burger, Angela Colbers, Hylke Waalewijn, Lisanne Bevers, Shaghayegh Mohsenian-Naghani, Anne Kamphuis

PK sub-studies – University of Cape Town: Helen McIlleron, Jennifer Norman, Lubbe Wiesner, Roeland Wasmann, Paolo Denti, Lufina Tsirizani Galileya

Toxicity sub-study: Eva Natukunda, Victor Musiime, Phillipa Musoke

Health Economics sub-study – University of York: Paul Revill, Simon Walker, Yingying Zhang Trial Committees

Independent Trial Steering Committee Members: Adeodata Kekitiinwa, Angela Mushavi, Febby Banda Kawamya, Denis Tindyebwa, Hermione Lyall, Ian Weller

Independent Data Monitoring Committee Members: Tim Peto, Philippa Musoke, Margaret Siwale, Rose Kambarami Funders

EDCTP: Johanna Roth, Pauline Beattie

Janssen Pharmaceuticals

Gilead Sciences.

Drug Contributors

Gilead Sciences Ltd., Janssen Pharmaceuticals, Viiv Healthcare, GSK Ltd., Cipla Ltd.

## Ethical approval

The trial was approved by ethics committees in Uganda (Joint Research Ethics Committee (JREC)), Zambia (University of Zambia Biomedical Research Ethics Committee (UNZABREC)), Zimbabwe (Joint Research Ethics Committee University of Zimbabwe College of Health Sciences (JREC), Research Council Zimbabwe (RCZ)), South Africa (University of Cape Town Human Research Ethics Committee) and United Kingdom (UCL Research Ethics Committee)

