## Supplemental File 1 for "CHAPAS-4 trial: second-line anchor drugs for children with HIV in Africa"

**Table S1: Additional baseline characteristics**

| | LPV/r N=227 | ATV/r N=231 | DRV/r N=232 | DTG N=229 | Total N=919 |
|---|---|---|---|---|---|
| Centre | | | | | |
| Uganda/Kampala | 49 (21.6%) | 51 (22.1%) | 51 (22.0%) | 50 (21.8%) | 201 (21.9%) |
| Uganda/Mbarara | 49 (21.6%) | 49 (21.2%) | 49 (21.1%) | 49 (21.4%) | 196 (21.3%) |
| Zambia/Lusaka | 30 (13.2%) | 31 (13.4%) | 29 (12.5%) | 31 (13.5%) | 121 (13.2%) |
| Zambia/Ndola | 18 (7.9%) | 18 (7.8%) | 19 (8.2%) | 19 (8.3%) | 74 (8.1%) |
| Zimbabwe/Harare | 54 (23.8%) | 54 (23.4%) | 56 (24.1%) | 55 (24.0%) | 219 (23.8%) |
| Zimbabwe/Bulawayo | 27 (11.9%) | 28 (12.1%) | 28 (12.1%) | 25 (10.9%) | 108 (11.8%) |

Values are n (%) or median (IQR)

ATV/r denotes ritonavir-boosted atazanavir, DRV/r ritonavir-boosted darunavir, DTG dolutegravir and LPV/r ritonavir-boosted lopinavir

**Table S2: VL comparisons at different thresholds and weeks**

| Comparison | Week 48 (n=907): difference (%) [95% CI] | p | Week 96 (n=908): difference (%) [95% CI] | p | Week 144 (n=488): difference (%) [95% CI] | p |
|---|---|---|---|---|---|---|
| **<400 copies/ml** | | | | | | |
| ATV/r vs. LPV/r | 5.8 [-1.0, 12.6] | 0.09 | 3.4 [-3.4, 10.2] | 0.33 | 1.5 [-7.8, 10.7] | 0.76 |
| DTG vs. LPV/r or ATV/r | 10.6 [6.0, 15.2] | <0.0001 | 9.7 [4.8, 14.5] | <0.0001 | 7.4 [0.4, 14.4] | 0.04 |
| DRV/r vs. LPV/r or ATV/r | 3.2 [-2.3, 8.8] | 0.25 | 5.6 [0.3, 11.0] | 0.04 | 4.3 [-3.2, 11.8] | 0.26 |
| DTG vs. DRV/r | 7.3 [2.0, 12.7] | 0.007 | 4.0 [-1.3, 9.4] | 0.14 | 3.1 [-4.7, 11.0] | 0.43 |
| **<60 copies/ml** | | | | | | |
| ATV/r vs. LPV/r | 4.8 [-3.1, 12.7] | 0.23 | 5.4 [-2.5, 13.2] | 0.18 | 6.1 [-5.0, 17.2] | 0.28 |
| DTG vs. LPV/r or ATV/r | 9.9 [3.8, 16.0] | 0.002 | 10.5 [4.4, 16.6] | 0.0007 | 8.1 [-0.8, 16.9] | 0.07 |
| DRV/r vs. LPV/r or ATV/r | 1.4 [-5.4, 8.1] | 0.69 | 3.1 [-3.5, 9.8] | 0.35 | 4.7 [-4.3, 13.8] | 0.31 |
| DTG vs. DRV/r | 8.5 [1.4, 15.7] | 0.02 | 7.4 [0.4, 14.4] | 0.04 | 3.3 [-6.4, 12.9] | 0.51 |
| **<1000 copies/ml** | | | | | | |
| ATV/r vs. LPV/r | 3.8 [-2.2, 9.9] | 0.22 | 2.9 [-3.5, 9.3] | 0.37 | 3.4 [-5.1, 12.0] | 0.43 |
| DTG vs. LPV/r or ATV/r | 8.0 [3.8, 12.1] | 0.0002 | 8.5 [4.1, 13.0] | 0.0002 | 5.0 [-1.4, 11.5] | 0.13 |
| DRV/r vs. LPV/r or ATV/r | 1.0 [-4.2, 6.1] | 0.71 | 4.9 [0.0, 9.9] | 0.048 | 4.6 [-2.0, 11.3] | 0.17 |
| DTG vs. DRV/r | 7.0 [2.0, 11.9] | 0.006 | 3.6 [-1.2, 8.4] | 0.15 | 0.2 [-6.7, 7.1] | 0.96 |

ATV/r denotes ritonavir-boosted atazanavir, DRV/r ritonavir-boosted darunavir, DTG dolutegravir, LPV/r ritonavir-boosted lopinavir and VL HIV viral load

**Table S3: Number of disease progression events of each type**

| | LPV/r | ATV/r | DRV/r | DTG |
|---|---|---|---|---|
| WHO 3 | 0 (0.0%) 0 | 0 (0.0%) 0 | 0 (0.0%) 0 | 1 (0.4%) 1 |
| WHO 4 | 2 (0.9%) 2 | 1 (0.4%) 1 | 2 (0.9%) 2 | 3 (1.3%) 3 |
| Death | 0 (0.0%) | 0 (0.0%) | 0 (0.0%) | 1 (0.4%) |
| Any | 2 (0.9%) 2 | 1 (0.4%) 1 | 2 (0.9%) 2 | 4 (1.7%) 5 |

Showing Number of patients with one or more event (% of patients) number of events
e.g., '2 (20.0%) 3,' would indicate a total of 3 events in a total of 2 patients
Corresponding hazard ratios and 95% confidence intervals:
ATV/r v LPV/r: 0.49 [0.04, 5.39]; DRV/r v LPV/r: 0.98 [0.14, 6.93]; DTG v LPV/r: 1.99 [0.36, 10.87]

ATV/r denotes ritonavir-boosted atazanavir, DRV/r ritonavir-boosted darunavir, DTG dolutegravir and LPV/r ritonavir-boosted lopinavir

**Table S4: Grade 3 and 4 adverse events during 96-week follow-up**

| | LPV/r N=227 | ATV/r N=231 | DRV/r N=232 | DTG N=229 | Total N=919 | p* |
|---|---|---|---|---|---|---|
| **Any** | **26 (11.5%) 36** | **69 (29.9%) 92** | **20 (8.6%) 28** | **12 (5.2%) 20** | **127 (13.8%) 176** | **<0.0001** |
| **CNS** | **0 (0.0%) 0** | **0 (0.0%) 0** | **1 (0.4%) 3** | **0 (0.0%) 0** | **1 (0.1%) 3** | **1.00** |
| **Psychiatric** | **3 (1.3%) 3** | **1 (0.4%) 1** | **1 (0.4%) 1** | **0 (0.0%) 0** | **5 (0.5%) 5** | **0.23** |
| **Lower Respiratory Tract** | **2 (0.9%) 2** | **0 (0.0%) 0** | **2 (0.9%) 2** | **1 (0.4%) 2** | **5 (0.5%) 6** | **0.53** |
| **Cardiovascular** | **0 (0.0%) 0** | **0 (0.0%) 0** | **0 (0.0%) 0** | **1 (0.4%) 1** | **1 (0.1%) 1** | **0.50** |
| **Eye** | **0 (0.0%) 0** | **1 (0.4%) 1** | **0 (0.0%) 0** | **0 (0.0%) 0** | **1 (0.1%) 1** | **0.75** |
| **Gastrointestinal** | **1 (0.4%) 1** | **1 (0.4%) 1** | **0 (0.0%) 0** | **0 (0.0%) 0** | **2 (0.2%) 2** | **0.50** |
| **Hepatic** | **2 (0.9%) 2** | **3 (1.3%) 3** | **1 (0.4%) 1** | **0 (0.0%) 0** | **6 (0.7%) 6** | **0.38** |
| **Musculoskeletal** | **0 (0.0%) 0** | **0 (0.0%) 0** | **0 (0.0%) 0** | **1 (0.4%) 1** | **1 (0.1%) 1** | **0.50** |
| **Skin** | **3 (1.3%) 5** | **1 (0.4%) 1** | **1 (0.4%) 1** | **0 (0.0%) 0** | **5 (0.5%) 7** | **0.23** |
| **Haematological** | **13 (5.7%) 14** | **9 (3.9%) 13** | **13 (5.6%) 17** | **8 (3.5%) 9** | **43 (4.7%) 53** | **0.58** |
| Pancytopenia, bone marrow depression | 0 (0.0%) 0 | 1 (0.4%) 2 | 0 (0.0%) 0 | 0 (0.0%) 0 | 1 (0.1%) 2 | |
| Anaemia with clinical symptoms | 3 (1.3%) 3 | 0 (0.0%) 0 | 4 (1.7%) 4 | 0 (0.0%) 0 | 7 (0.8%) 7 | |
| Thrombocytopenia | 3 (1.3%) 3 | 0 (0.0%) 0 | 2 (0.9%) 4 | 2 (0.9%) 2 | 7 (0.8%) 9 | |
| Leucopenia | 0 (0.0%) 0 | 1 (0.4%) 1 | 0 (0.0%) 0 | 0 (0.0%) 0 | 1 (0.1%) 1 | |
| Neutropenia | 3 (1.3%) 3 | 6 (2.6%) 6 | 4 (1.7%) 5 | 2 (0.9%) 2 | 15 (1.6%) 16 | |
| Anaemia with no clinical symptoms | 4 (1.8%) 4 | 2 (0.9%) 2 | 3 (1.3%) 3 | 4 (1.7%) 4 | 13 (1.4%) 13 | |
| Lymphopenia | 1 (0.4%) 1 | 2 (0.9%) 2 | 1 (0.4%) 1 | 1 (0.4%) 1 | 5 (0.5%) 5 | |
| **Biochemical** | **3 (1.3%) 3** | **60 (26.0%) 70** | **1 (0.4%) 1** | **0 (0.0%) 0** | **64 (7.0%) 74** | **<0.0001** |
| Raised liver enzymes | 0 (0.0%) 0 | 1 (0.4%) 1 | 0 (0.0%) 0 | 0 (0.0%) 0 | 1 (0.1%) 1 | |
| Raised AST | 1 (0.4%) 1 | 2 (0.9%) 2 | 0 (0.0%) 0 | 0 (0.0%) 0 | 3 (0.3%) 3 | |
| Raised ALT | 1 (0.4%) 1 | 1 (0.4%) 1 | 0 (0.0%) 0 | 0 (0.0%) 0 | 2 (0.2%) 2 | |
| Raised bilirubin | 1 (0.4%) 1 | 57 (24.7%) 66 | 1 (0.4%) 1 | 0 (0.0%) 0 | 59 (6.4%) 68 | |
| **Systemic** | **1 (0.4%) 1** | **0 (0.0%) 0** | **0 (0.0%) 0** | **2 (0.9%) 2** | **3 (0.3%) 3** | **0.20** |
| **Specific Infections** | **3 (1.3%) 3** | **0 (0.0%) 0** | **1 (0.4%) 1** | **4 (1.7%) 4** | **8 (0.9%) 8** | **0.11** |
| **Undiagnosed Fevers** | **0 (0.0%) 0** | **1 (0.4%) 1** | **0 (0.0%) 0** | **0 (0.0%) 0** | **1 (0.1%) 1** | **0.75** |
| **Tumours** | **0 (0.0%) 0** | **0 (0.0%) 0** | **0 (0.0%) 0** | **1 (0.4%) 1** | **1 (0.1%) 1** | **0.50** |
| **Pregnancy Associated** | **1 (0.4%) 1** | **0 (0.0%) 0** | **0 (0.0%) 0** | **0 (0.0%) 0** | **1 (0.1%) 1** | **0.25** |
| **Other** | **1 (0.4%) 1** | **1 (0.4%) 1** | **1 (0.4%) 1** | **0 (0.0%) 0** | **3 (0.3%) 3** | **0.90** |

Excluding extended follow-up after 96 weeks

Detail within body system provided where p≤0.05 or ≥10% of children experienced an event

Showing number of patients with one or more event (% of patients) number of events
*Fisher's exact test

ALT denotes alanine aminotransferase, AST aspartate aminotransaminase, ATV/r denotes ritonavir-boosted atazanavir, CNS central nervous system, DRV/r ritonavir-boosted darunavir, DTG dolutegravir and LPV/r ritonavir-boosted lopinavir

**Table S5: SAEs during 96-week follow-up**

| | LPV/r N=227 | ATV/r N=231 | DRV/r N=232 | DTG N=229 | Total N=919 | p* |
|---|---|---|---|---|---|---|
| **Any** | **10 (4.4%) 10** | **5 (2.2%) 6** | **8 (3.4%) 9** | **6 (2.6%) 6** | **29 (3.2%) 31** | **0.55** |
| CNS | 0 (0.0%) 0 | 0 (0.0%) 0 | 1 (0.4%) 2 | 0 (0.0%) 0 | 1 (0.1%) 2 | 1.00 |
| Upper Respiratory Tract | 1 (0.4%) 1 | 1 (0.4%) 1 | 0 (0.0%) 0 | 0 (0.0%) 0 | 2 (0.2%) 2 | 0.50 |
| Lower Respiratory Tract | 3 (1.3%) 3 | 0 (0.0%) 0 | 2 (0.9%) 2 | 1 (0.4%) 1 | 6 (0.7%) 6 | 0.26 |
| Gastrointestinal | 1 (0.4%) 1 | 1 (0.4%) 1 | 0 (0.0%) 0 | 0 (0.0%) 0 | 2 (0.2%) 2 | 0.50 |
| Hepatic | 0 (0.0%) 0 | 1 (0.4%) 1 | 0 (0.0%) 0 | 0 (0.0%) 0 | 1 (0.1%) 1 | 0.75 |
| Skin | 2 (0.9%) 2 | 0 (0.0%) 0 | 1 (0.4%) 1 | 0 (0.0%) 0 | 3 (0.3%) 3 | 0.25 |
| Haematological | 1 (0.4%) 1 | 1 (0.4%) 2 | 2 (0.9%) 2 | 0 (0.0%) 0 | 4 (0.4%) 5 | 0.81 |
| Systemic | 0 (0.0%) 0 | 0 (0.0%) 0 | 0 (0.0%) 0 | 1 (0.4%) 1 | 1 (0.1%) 1 | 0.50 |
| Specific Infections | 2 (0.9%) 2 | 0 (0.0%) 0 | 1 (0.4%) 1 | 4 (1.7%) 4 | 7 (0.8%) 7 | 0.12 |
| Undiagnosed Fevers | 0 (0.0%) 0 | 1 (0.4%) 1 | 0 (0.0%) 0 | 0 (0.0%) 0 | 1 (0.1%) 1 | 0.75 |
| Other | 0 (0.0%) 0 | 0 (0.0%) 0 | 1 (0.4%) 1 | 0 (0.0%) 0 | 1 (0.1%) 1 | 1.00 |

Excluding extended follow-up after 96 weeks

Detail within body system provided where p≤0.05 or ≥10% of children experienced an event

Showing number of patients with one or more episode (% of patients) number of episodes

*Fisher's exact test

ATV/r denotes ritonavir-boosted atazanavir, DRV/r ritonavir-boosted darunavir, DTG dolutegravir and LPV/r ritonavir-boosted lopinavir

**Table S6: Weight-for-age at week 96 by weight-for-age at week 0 by anchor drug**

**LPV/r**

| Week 0 | <-3 N=23 | -3 to <-2 N=49 | -2 to <-1 N=71 | -1 to <0 N=55 | 0 to <1 N=16 | 1 to <2 N=1 | Total N=215 |
|---|---|---|---|---|---|---|---|
| **Week 96** | | | | | | | |
| <-3 | 15 (65.2%) | 5 (10.2%) | 1 (1.4%) | 0 (0.0%) | 0 (0.0%) | 0 (0.0%) | 21 (9.8%) |
| -3 to <-2 | 8 (34.8%) | 34 (69.4%) | 7 (9.9%) | 0 (0.0%) | 0 (0.0%) | 0 (0.0%) | 49 (22.8%) |
| -2 to <-1 | 0 (0.0%) | 9 (18.4%) | 51 (71.8%) | 14 (25.5%) | 0 (0.0%) | 0 (0.0%) | 74 (34.4%) |
| -1 to <0 | 0 (0.0%) | 1 (2.0%) | 12 (16.9%) | 36 (65.5%) | 5 (31.3%) | 0 (0.0%) | 54 (25.1%) |
| 0 to <1 | 0 (0.0%) | 0 (0.0%) | 0 (0.0%) | 5 (9.1%) | 11 (68.8%) | 0 (0.0%) | 16 (7.4%) |
| 1 to <2 | 0 (0.0%) | 0 (0.0%) | 0 (0.0%) | 0 (0.0%) | 0 (0.0%) | 1 (100.0%) | 1 (0.5%) |

**ATV/r**

| Week 0 | <-3 N=31 | -3 to <-2 N=50 | -2 to <-1 N=82 | -1 to <0 N=42 | 0 to <1 N=13 | 1 to <2 N=4 | Total N=222 |
|---|---|---|---|---|---|---|---|
| **Week 96** | | | | | | | |
| <-3 | 18 (58.1%) | 2 (4.0%) | 0 (0.0%) | 0 (0.0%) | 0 (0.0%) | 0 (0.0%) | 20 (9.0%) |
| -3 to <-2 | 11 (35.5%) | 26 (52.0%) | 5 (6.1%) | 0 (0.0%) | 0 (0.0%) | 0 (0.0%) | 42 (18.9%) |
| -2 to <-1 | 2 (6.5%) | 21 (42.0%) | 57 (69.5%) | 1 (2.4%) | 0 (0.0%) | 1 (25.0%) | 82 (36.9%) |
| -1 to <0 | 0 (0.0%) | 1 (2.0%) | 19 (23.2%) | 33 (78.6%) | 0 (0.0%) | 0 (0.0%) | 53 (23.9%) |
| 0 to <1 | 0 (0.0%) | 0 (0.0%) | 1 (1.2%) | 7 (16.7%) | 10 (76.9%) | 1 (25.0%) | 19 (8.6%) |
| 1 to <2 | 0 (0.0%) | 0 (0.0%) | 0 (0.0%) | 1 (2.4%) | 2 (15.4%) | 2 (50.0%) | 5 (2.3%) |
| 2 to <3 | 0 (0.0%) | 0 (0.0%) | 0 (0.0%) | 0 (0.0%) | 1 (7.7%) | 0 (0.0%) | 1 (0.5%) |

**DRV/r**

| Week 0 | <-3 N=24 | -3 to <-2 N=63 | -2 to <-1 N=75 | -1 to <0 N=44 | 0 to <1 N=18 | 1 to <2 N=1 | Total N=225 |
|---|---|---|---|---|---|---|---|
| **Week 96** | | | | | | | |
| <-3 | 11 (45.8%) | 3 (4.8%) | 0 (0.0%) | 0 (0.0%) | 0 (0.0%) | 0 (0.0%) | 14 (6.2%) |
| -3 to <-2 | 12 (50.0%) | 34 (54.0%) | 12 (16.0%) | 0 (0.0%) | 0 (0.0%) | 0 (0.0%) | 58 (25.8%) |
| -2 to <-1 | 1 (4.2%) | 20 (31.7%) | 48 (64.0%) | 5 (11.4%) | 0 (0.0%) | 0 (0.0%) | 74 (32.9%) |
| -1 to <0 | 0 (0.0%) | 6 (9.5%) | 14 (18.7%) | 36 (81.8%) | 4 (22.2%) | 0 (0.0%) | 60 (26.7%) |
| 0 to <1 | 0 (0.0%) | 0 (0.0%) | 0 (0.0%) | 3 (6.8%) | 10 (55.6%) | 0 (0.0%) | 13 (5.8%) |
| 1 to <2 | 0 (0.0%) | 0 (0.0%) | 1 (1.3%) | 0 (0.0%) | 4 (22.2%) | 1 (100.0%) | 6 (2.7%) |

**DTG**

| Week 0 | <-3 N=26 | -3 to <-2 N=61 | -2 to <-1 N=73 | -1 to <0 N=53 | 0 to <1 N=8 | 1 to <2 N=1 | Total N=222 |
|---|---|---|---|---|---|---|---|
| **Week 96** | | | | | | | |
| <-3 | 14 (53.8%) | 3 (4.9%) | 0 (0.0%) | 0 (0.0%) | 0 (0.0%) | 0 (0.0%) | 17 (7.7%) |
| -3 to <-2 | 11 (42.3%) | 30 (49.2%) | 6 (8.2%) | 0 (0.0%) | 0 (0.0%) | 0 (0.0%) | 47 (21.2%) |
| -2 to <-1 | 1 (3.8%) | 25 (41.0%) | 42 (57.5%) | 10 (18.9%) | 0 (0.0%) | 0 (0.0%) | 78 (35.1%) |
| -1 to <0 | 0 (0.0%) | 3 (4.9%) | 22 (30.1%) | 34 (64.2%) | 3 (37.5%) | 0 (0.0%) | 62 (27.9%) |
| 0 to <1 | 0 (0.0%) | 0 (0.0%) | 2 (2.7%) | 7 (13.2%) | 4 (50.0%) | 0 (0.0%) | 13 (5.9%) |
| 1 to <2 | 0 (0.0%) | 0 (0.0%) | 1 (1.4%) | 2 (3.8%) | 1 (12.5%) | 1 (100.0%) | 5 (2.3%) |

ATV/r denotes ritonavir-boosted atazanavir, DRV/r ritonavir-boosted darunavir, DTG dolutegravir and LPV/r ritonavir-boosted lopinavir

**Figure S1: Treatment received over time (extended follow-up from weeks 120-168)**

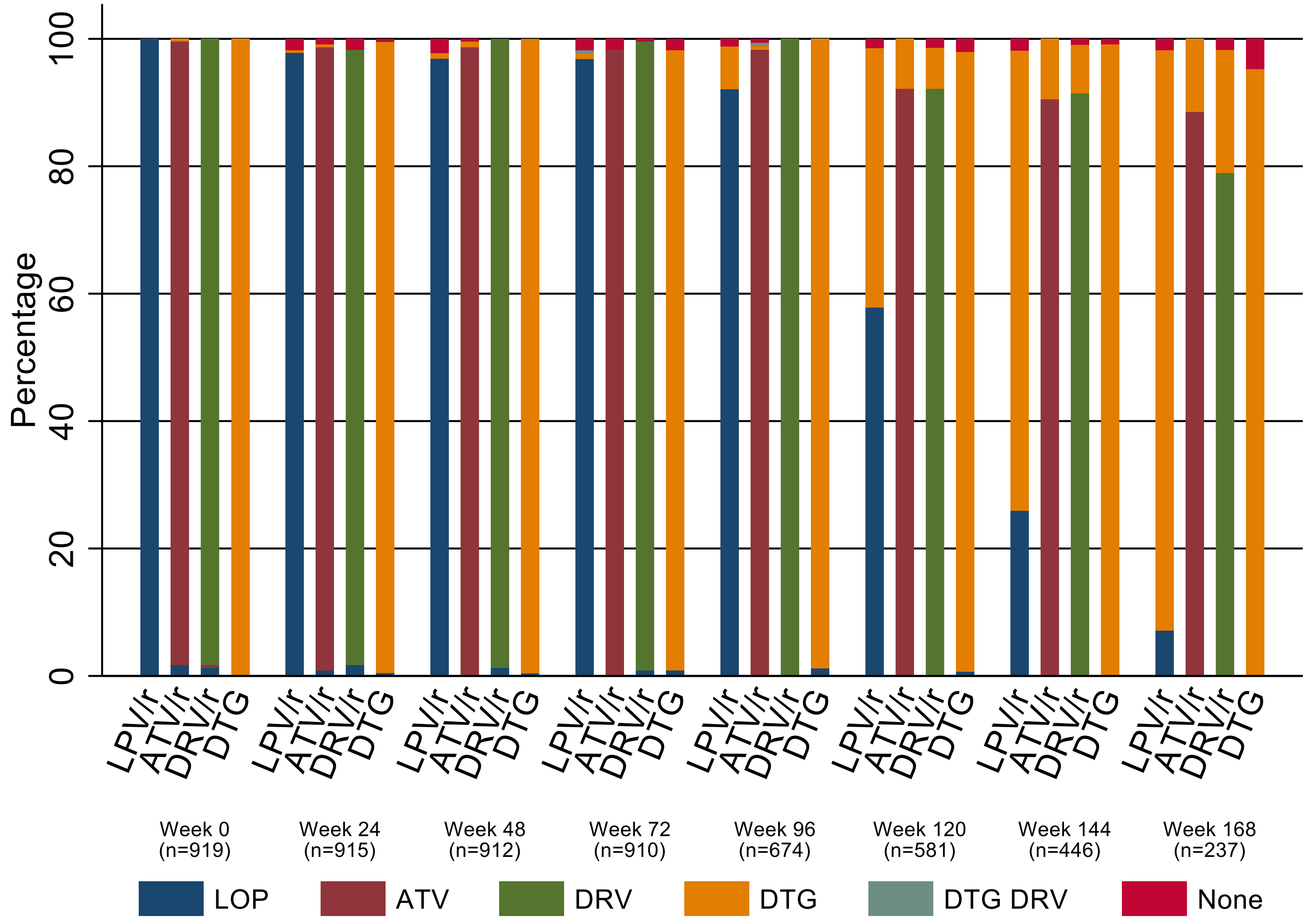

ATV/r denotes ritonavir-boosted atazanavir, DRV/r ritonavir-boosted darunavir, DTG dolutegravir and LPV/r ritonavir-boosted lopinavir

**Figure S2: Subgroup analyses for primary endpoint VL <400 copies/ml at 96 weeks: ATV/r vs. LPV/r**

| | LPV/r | ATV/r | ATV/r-LPV/r difference (%) [95% CI] | Interaction p |
|---|---|---|---|---|
| **01 PARTICIPANT HAS BEEN RANDOMISED TO: NRTI BACKBONE RANDOMISATION** | | | | |
| Standard-of-care | 83/112(74.1%) | 91/113(80.5%) | 4.9 [-3.1, 12.9] | 0.54 |
| TAF | 97/111(87.4%) | 102/116(87.9%) | 0.7 [-10.3, 11.7] | |
| **02 SITE** | | | | |
| Kampala | 46/48(95.8%) | 47/51(92.2%) | -10.7 [-42.0, 20.6] | 0.32 |
| Mbarara | 40/48(83.3%) | 40/49(81.6%) | -1.7 [-16.8, 13.4] | |
| Lusaka | 19/29(65.5%) | 25/30(83.3%) | 10.3 [0.5, 20.1] | |
| Ndola | 11/18(61.1%) | 15/18(83.3%) | 11.5 [1.0, 21.9] | |
| Harare | 44/53(83.0%) | 42/53(79.2%) | -3.6 [-18.4, 11.2] | |
| Bulawayo | 20/27(74.1%) | 24/28(85.7%) | 8.4 [-3.9, 20.6] | |
| **03 FAILING FIRST-LINE NRTI** | | | | |
| ABC | 88/118(74.6%) | 100/123(81.3%) | 5.3 [-2.6, 13.1] | 0.46 |
| ZDV | 92/105(87.6%) | 93/106(87.7%) | 0.2 [-11.4, 11.7] | |
| **04 VL AT TRIAL ENTRY (COPIES/ML)** | | | | |
| <10000 | 70/85(82.4%) | 70/86(81.4%) | -0.9 [-12.4, 10.5] | 0.47 |
| 10000-99999 | 84/105(80.0%) | 94/107(87.9%) | 7.6 [-1.0, 16.2] | |
| >=100000 | 26/33(78.8%) | 29/36(80.6%) | 1.5 [-14.3, 17.4] | |
| **05 COUNTRY** | | | | |
| Uganda | 86/96(89.6%) | 87/100(87.0%) | -3.6 [-16.9, 9.6] | 0.13 |
| Zambia | 30/47(63.8%) | 40/48(83.3%) | 11.3 [3.3, 19.3] | |
| Zimbabwe | 64/80(80.0%) | 66/81(81.5%) | 1.3 [-9.2, 11.8] | |

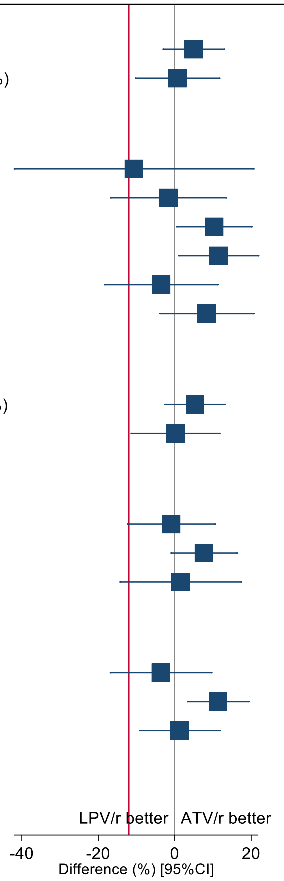

Red line denotes non-inferiority margin (12%)

| | LPV/r | ATV/r | ATV/r-LPV/r difference (%) [95% CI] | Interaction p |
|---|---|---|---|---|
| **07 SEX** | | | | |
| Male | 92/116(79.3%) | 106/128(82.8%) | 3.2 [-5.5, 11.9] | 0.90 |
| Female | 88/107(82.2%) | 87/101(86.1%) | 4.0 [-5.6, 13.6] | |
| **08 WEIGHT (KG)** | | | | |
| <20 | 46/51(90.2%) | 41/48(85.4%) | -6.9 [-27.5, 13.6] | 0.22 |
| 20-<25 | 44/54(81.5%) | 57/63(90.5%) | 8.9 [-1.3, 19.2] | |
| 25-<35 | 60/71(84.5%) | 65/80(81.3%) | -3.3 [-16.2, 9.5] | |
| >=35 | 30/47(63.8%) | 30/38(78.9%) | 8.7 [-0.5, 17.8] | |
| **09 FAILING FIRST LINE NNRTI** | | | | |
| EFV | 96/129(74.4%) | 104/127(81.9%) | 5.9 [-1.7, 13.5] | 0.23 |
| NVP | 84/94(89.4%) | 89/102(87.3%) | -3.0 [-16.2, 10.3] | |
| **10 WEIGHT-FOR-AGE Z-SCORE*** | | | | |
| <=-2.1 | 51/66(77.3%) | 58/75(77.3%) | 0.0 [-11.1, 11.2] | 0.60 |
| >-2.1 to <=-1.13 | 59/75(78.7%) | 67/80(83.8%) | 4.4 [-5.6, 14.5] | |
| >-1.13 | 70/82(85.4%) | 68/74(91.9%) | 8.0 [-2.4, 18.4] | |
| **11 CD4 AT TRIAL ENTRY (CELLS/MM^3)*** | | | | |
| <=499 | 51/72(70.8%) | 54/67(80.6%) | 6.9 [-2.1, 15.8] | 0.50 |
| >499 - <=852 | 57/70(81.4%) | 71/84(84.5%) | 3.0 [-8.1, 14.1] | |
| >852 | 70/79(88.6%) | 63/73(86.3%) | -3.1 [-18.1, 11.8] | |

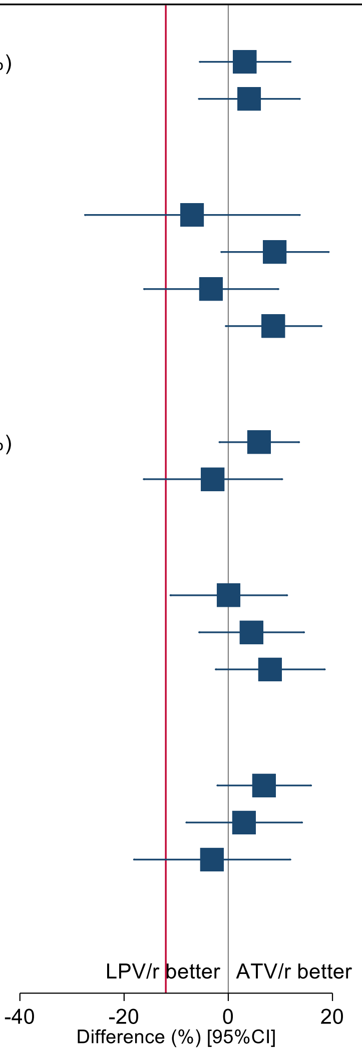

Red line denotes non-inferiority margin (12%)
*Terciles
AGE AT LAST BIRTHDAY (YEARS) (model not fitted due to low numbers) (ATV/r vs. LPV/r):
3-4: 12/14 (85.7%) vs. 11/11 (100.0%); 5-9: 72/82 (87.8%) vs. 81/92 (88.0%); 10-15: 109/133 (82.0%) vs. 88/120 (73.3%)

ABC denotes abacavir, ATV/r ritonavir-boosted atazanavir, EFV efavirenz, LPV/r ritonavir-boosted lopinavir, NNRTI non-nucleoside reverse transcriptase inhibitor, NRTI nucleoside/nucleotide reverse transcriptase inhibitor, NVP nevirapine, TAF tenofovir alafenamide fumarate, VL HIV viral load, ZDV zidovudine

### Figure S3: subgroup analyses for primary endpoint VL <400 copies/ml at 96 weeks: DRV/r vs. ATV/r and LPV/r combined

| | LPV/r / ATV/r | DRV/r | DRV/r-LPV/r / ATV/r difference (%) [95% CI] | Interaction p |
|---|---|---|---|---|
| **01 PARTICIPANT HAS BEEN RANDOMISED TO: NRTI BACKBONE RANDOMISATION** | | | | |
| Standard-of-care | 174/225(77.3%) | 98/113(86.7%) | 7.3 [1.2, 13.3] | 0.36 |
| TAF | 199/227(87.7%) | 105/117(89.7%) | 2.5 [-5.8, 10.9] | |
| **02 SITE** | | | | |
| Kampala | 93/99(93.9%) | 50/51(98.0%) | 10.2 [-1.5, 21.9] | 0.98 |
| Mbarara | 80/97(82.5%) | 42/48(87.5%) | 4.5 [-5.6, 14.6] | |
| Lusaka | 44/59(74.6%) | 24/29(82.8%) | 5.4 [-5.1, 15.9] | |
| Ndola | 26/36(72.2%) | 14/18(77.8%) | 3.4 [-10.5, 17.4] | |
| Harare | 86/106(81.1%) | 48/56(85.7%) | 3.9 [-5.6, 13.3] | |
| Bulawayo | 44/55(80.0%) | 25/28(89.3%) | 7.4 [-3.3, 18.2] | |
| **03 FAILING FIRST-LINE NRTI** | | | | |
| ABC | 188/241(78.0%) | 105/122(86.1%) | 6.4 [0.3, 12.5] | 0.64 |
| ZDV | 185/211(87.7%) | 98/108(90.7%) | 3.8 [-4.6, 12.2] | |
| **04 VL AT TRIAL ENTRY (COPIES/ML)** | | | | |
| <10000 | 140/171(81.9%) | 59/71(83.1%) | 1.1 [-8.0, 10.2] | 0.30 |
| 10000-99999 | 178/212(84.0%) | 122/132(92.4%) | 9.0 [2.7, 15.3] | |
| >=100000 | 55/69(79.7%) | 22/27(81.5%) | 1.4 [-12.3, 15.1] | |
| **05 COUNTRY** | | | | |
| Uganda | 173/196(88.3%) | 92/99(92.9%) | 6.2 [-2.1, 14.4] | 0.97 |
| Zambia | 70/95(73.7%) | 38/47(80.9%) | 4.7 [-4.0, 13.3] | |
| Zimbabwe | 130/161(80.7%) | 73/84(86.9%) | 5.2 [-2.3, 12.8] | |

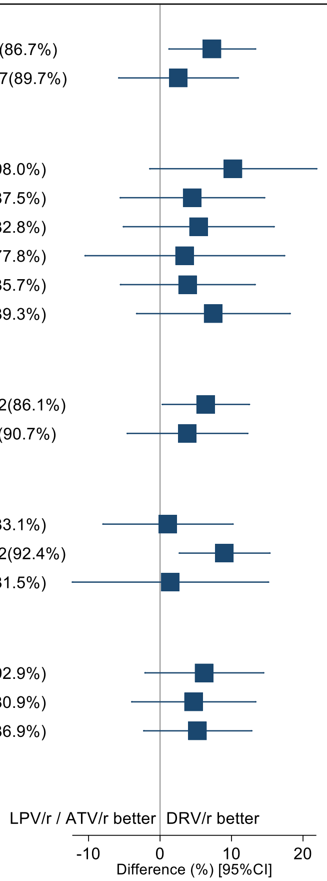

| | LPV/r / ATV/r | DRV/r | DRV/r-LPV/r / ATV/r difference (%) [95% CI] | Interaction p |
|---|---|---|---|---|
| **07 SEX** | | | | |
| Male | 198/244(81.1%) | 108/119(90.8%) | 8.9 [2.8, 14.9] | 0.14 |
| Female | 175/208(84.1%) | 95/111(85.6%) | 1.4 [-6.6, 9.4] | |
| **08 WEIGHT (KG)** | | | | |
| <20 | 87/99(87.9%) | 49/50(98.0%) | 13.4 [7.0, 19.9] | 0.38 |
| 20-<25 | 101/117(86.3%) | 46/52(88.5%) | 2.3 [-9.0, 13.7] | |
| 25-<35 | 125/151(82.8%) | 75/86(87.2%) | 4.1 [-4.0, 12.2] | |
| >=35 | 60/85(70.6%) | 33/42(78.6%) | 4.8 [-3.9, 13.4] | |
| **09 FAILING FIRST LINE NNRTI** | | | | |
| EFV | 200/256(78.1%) | 106/122(86.9%) | 7.0 [1.0, 13.0] | 0.36 |
| NVP | 173/196(88.3%) | 97/108(89.8%) | 2.0 [-7.1, 11.0] | |
| **10 WEIGHT-FOR-AGE Z-SCORE*** | | | | |
| <=-2.1 | 109/141(77.3%) | 68/77(88.3%) | 8.4 [1.7, 15.0] | 0.42 |
| >-2.1 to <=-1.13 | 126/155(81.3%) | 74/84(88.1%) | 6.0 [-1.5, 13.5] | |
| >-1.13 | 138/156(88.5%) | 61/69(88.4%) | -0.1 [-11.5, 11.4] | |
| **11 CD4 AT TRIAL ENTRY (CELLS/MM^3)*** | | | | |
| <=499 | 105/139(75.5%) | 63/78(80.8%) | 3.7 [-3.9, 11.3] | 0.68 |
| >499 - <=852 | 128/154(83.1%) | 67/73(91.8%) | 8.5 [1.0, 16.0] | |
| >852 | 133/152(87.5%) | 71/77(92.2%) | 5.9 [-3.3, 15.1] | |

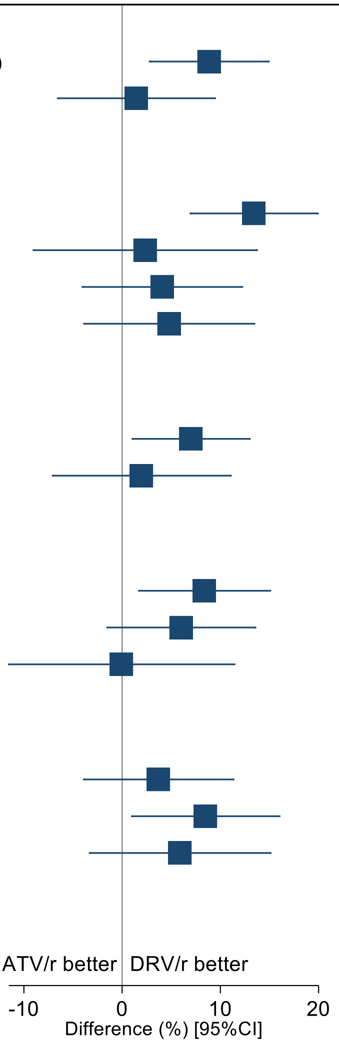

*Terciles
AGE AT LAST BIRTHDAY (YEARS) (model not fitted due to low numbers) (DRV/r vs. LPV/r / ATV/r):
3-4: 7/7 (100.0%) vs. 23/25 (92.0%); 5-9: 88/94 (93.6%) vs. 153/174 (87.9%); 10-15: 108/129 (83.7%) vs. 197/253 (77.9%)

ABC denotes abacavir, ATV/r ritonavir-boosted atazanavir, DRV/r ritonavir-boosted darunavir, EFV efavirenz, LPV/r ritonavir-boosted lopinavir, NNRTI non-nucleoside reverse transcriptase inhibitor, NRTI nucleoside/nucleotide reverse transcriptase inhibitor, NVP nevirapine, TAF tenofovir alafenamide fumarate, VL HIV viral load, ZDV zidovudine

### Figure S4: subgroup analyses for primary endpoint VL <400 copies/ml at 96 weeks: DTG vs. ATV/r and LPV/r combined

| | LPV/r / ATV/r | DTG | DTG-LPV/r / ATV/r difference (%) [95% CI] | Interaction p |
|---|---|---|---|---|
| **01 PARTICIPANT HAS BEEN RANDOMISED TO: NRTI BACKBONE RANDOMISATION** | | | | |
| Standard-of-care | 174/225(77.3%) | 106/116(91.4%) | 10.5 [5.4, 15.6] | 0.33 |
| TAF | 199/227(87.7%) | 102/110(92.7%) | 6.0 [-1.2, 13.2] | |
| **03 FAILING FIRST-LINE NRTI** | | | | |
| ABC | 188/241(78.0%) | 106/120(88.3%) | 7.7 [2.2, 13.2] | 0.41 |
| ZDV | 185/211(87.7%) | 102/106(96.2%) | 10.7 [4.8, 16.6] | |
| **04 VL AT TRIAL ENTRY (COPIES/ML)** | | | | |
| <10000 | 140/171(81.9%) | 77/83(92.8%) | 9.4 [3.4, 15.4] | 0.95 |
| 10000-99999 | 178/212(84.0%) | 98/106(92.5%) | 8.2 [2.1, 14.3] | |
| >=100000 | 55/69(79.7%) | 33/36(91.7%) | 8.9 [1.1, 16.7] | |
| **05 COUNTRY** | | | | |
| Uganda | 173/196(88.3%) | 94/98(95.9%) | 9.8 [3.3, 16.2] | 0.07 |
| Zambia | 70/95(73.7%) | 38/49(77.6%) | 2.3 [-6.1, 10.8] | |
| Zimbabwe | 130/161(80.7%) | 76/79(96.2%) | 12.9 [8.1, 17.7] | |

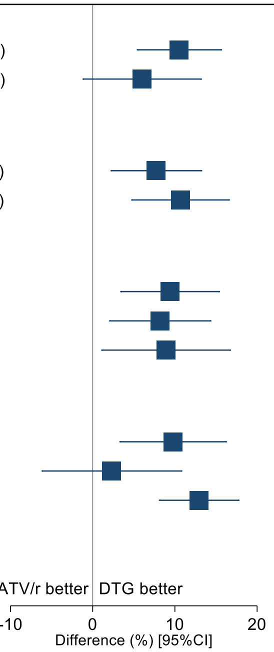

SITE (model not fitted due to low numbers) (DTG vs. LPV/r / ATV/r):
Kampala: 47/49 (95.9%) vs. 93/99 (93.9%); Mbarara: 47/49 (95.9%) vs. 80/97 (82.5%); Lusaka: 23/30 (76.7%) vs. 44/59 (74.6%); Ndola: 15/19 (78.9%) vs. 26/36 (72.2%); Harare 52/55 (94.5%) vs. 86/106 (81.1%); Bulawayo: 24/24 (100.0%) vs. 44/55 (80.0%)

| | LPV/r / ATV/r | DTG | DTG-LPV/r / ATV/r difference (%) [95% CI] | Interaction p |
|---|---|---|---|---|
| **06 AGE AT LAST BIRTHDAY (YEARS)** | | | | |
| 3-4 | 23/25(92.0%) | 4/6(66.7%) | -31.7 [-81.7, 18.3] | 0.07 |
| 5-9 | 153/174(87.9%) | 79/82(96.3%) | 10.5 [4.1, 16.8] | |
| 10-15 | 197/253(77.9%) | 125/138(90.6%) | 9.8 [4.7, 14.9] | |
| **07 SEX** | | | | |
| Male | 198/244(81.1%) | 115/125(92.0%) | 9.5 [4.0, 14.9] | 0.72 |
| Female | 175/208(84.1%) | 93/101(92.1%) | 7.7 [1.3, 14.1] | |
| **08 WEIGHT (KG)** | | | | |
| <20 | 87/99(87.9%) | 37/41(90.2%) | 2.7 [-9.5, 14.8] | 0.61 |
| 20-<25 | 101/117(86.3%) | 49/52(94.2%) | 8.4 [0.3, 16.5] | |
| 25-<35 | 125/151(82.8%) | 76/82(92.7%) | 8.9 [2.5, 15.2] | |
| >=35 | 60/85(70.6%) | 46/51(90.2%) | 10.9 [5.5, 16.4] | |
| **09 FAILING FIRST LINE NNRTI** | | | | |
| EFV | 200/256(78.1%) | 112/129(86.8%) | 6.5 [0.9, 12.0] | 0.02 |
| NVP | 173/196(88.3%) | 96/97(99.0%) | 14.4 [10.1, 18.6] | |
| **10 WEIGHT-FOR-AGE Z-SCORE*** | | | | |
| <=-2.1 | 109/141(77.3%) | 76/83(91.6%) | 10.3 [4.8, 15.8] | 0.48 |
| >-2.1 to <=-1.13 | 126/155(81.3%) | 64/69(92.8%) | 9.5 [3.4, 15.7] | |
| >-1.13 | 138/156(88.5%) | 68/74(91.9%) | 4.2 [-5.0, 13.4] | |
| **11 CD4 AT TRIAL ENTRY (CELLS/MM^3)*** | | | | |
| <=499 | 105/139(75.5%) | 73/80(91.3%) | 10.8 [5.4, 16.2] | 0.55 |
| >499 - <=852 | 128/154(83.1%) | 69/77(89.6%) | 5.8 [-1.7, 13.3] | |
| >852 | 133/152(87.5%) | 63/66(95.5%) | 9.5 [2.2, 16.9] | |

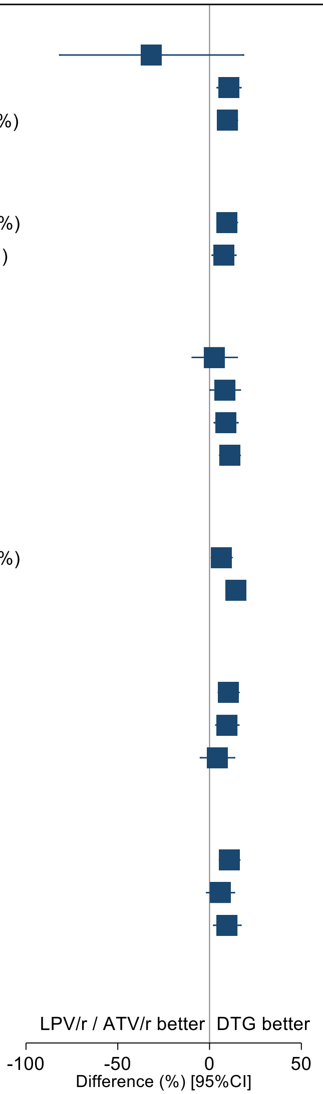

*Terciles

ABC denotes abacavir, ATV/r ritonavir-boosted atazanavir, DTG dolutegravir, EFV efavirenz, LPV/r ritonavir-boosted lopinavir, NNRTI non-nucleoside reverse transcriptase inhibitor, NRTI nucleoside/nucleotide reverse transcriptase inhibitor, NVP nevirapine, TAF tenofovir alafenamide fumarate, VL HIV viral load, ZDV zidovudine

**Figure S5: Changes over time in (a) CD4 and (b) CD4%**

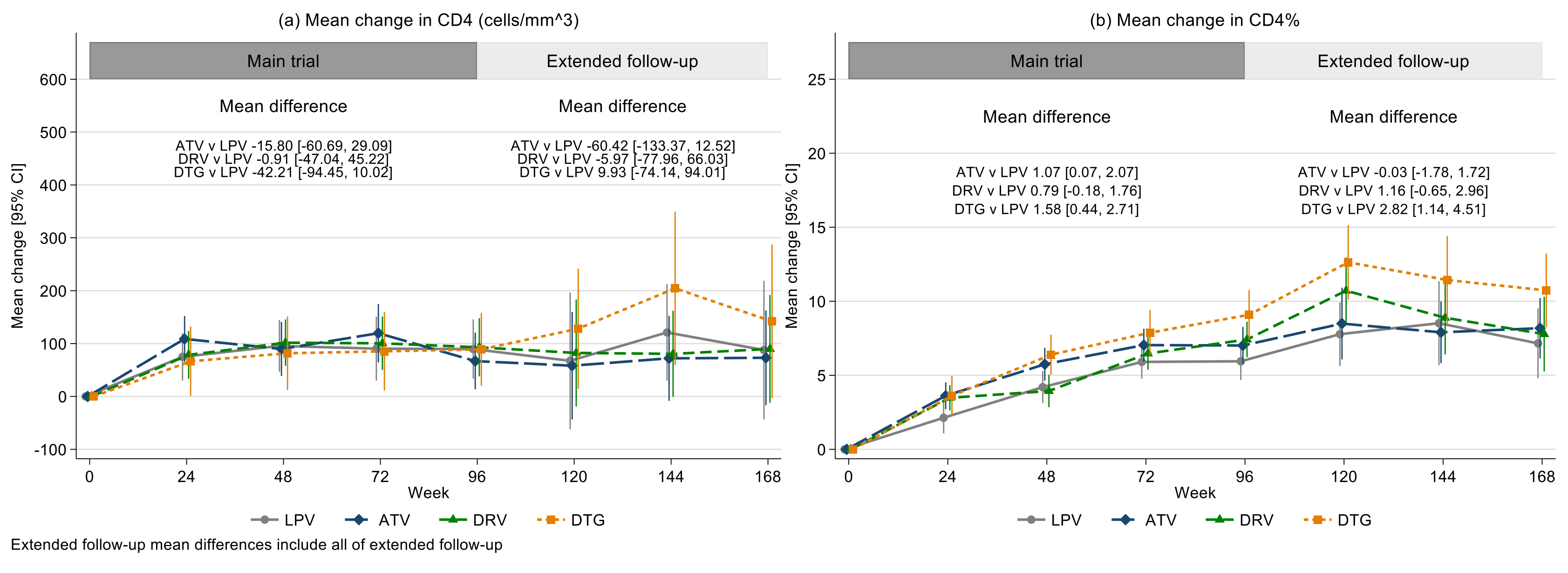

ATV denotes ritonavir-boosted atazanavir, DRV ritonavir-boosted darunavir, DTG dolutegravir and LPV ritonavir-boosted lopinavir

**Figure S6: Change in weight-for-age by combined NRTI backbone and anchor drug**

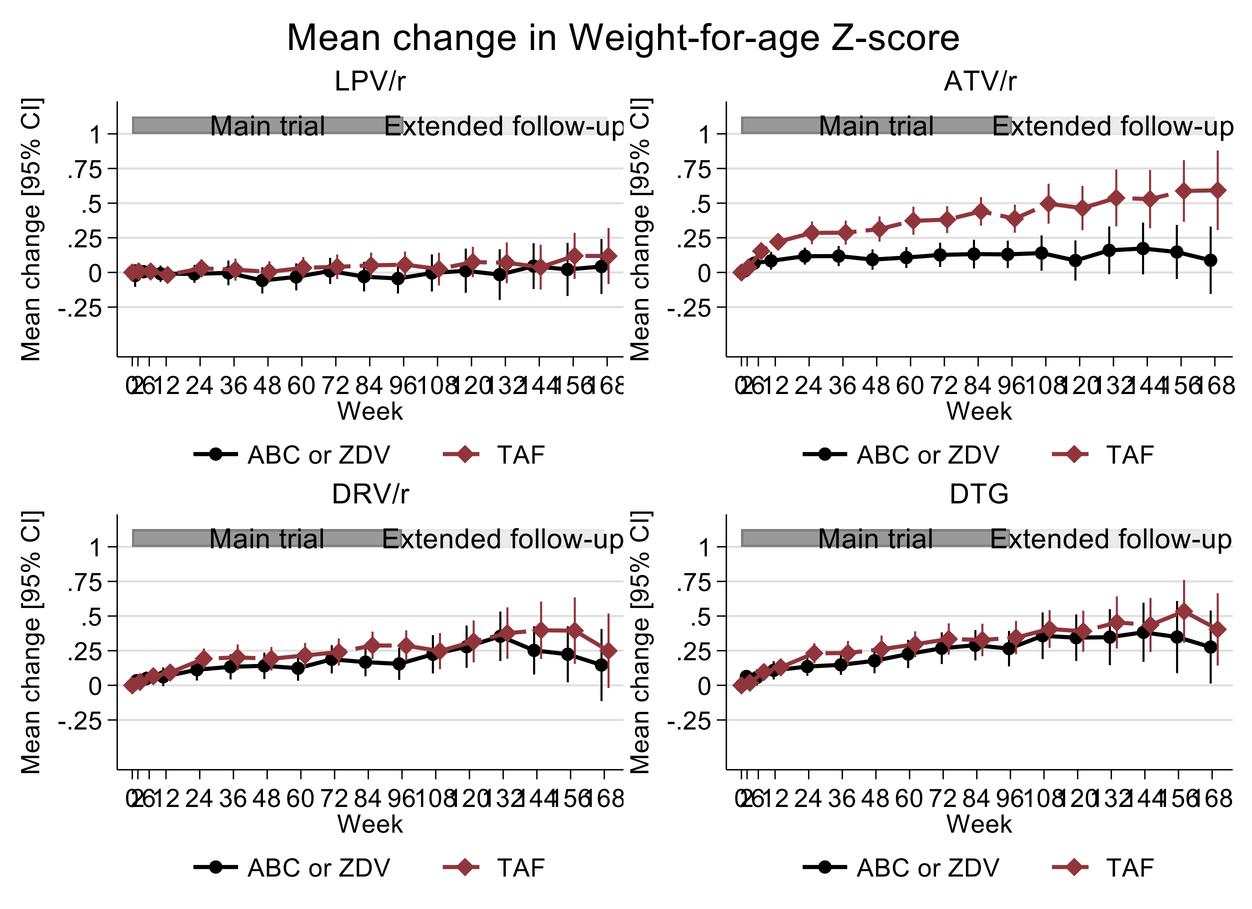

ABC denotes abacavir, ATV/r ritonavir-boosted atazanavir, DRV/r ritonavir-boosted darunavir, DTG dolutegravir, LPV/r ritonavir-boosted lopinavir, TAF tenofovir alafenamide fumarate and ZDV zidovudine

**Figure S7: Change in bilirubin**

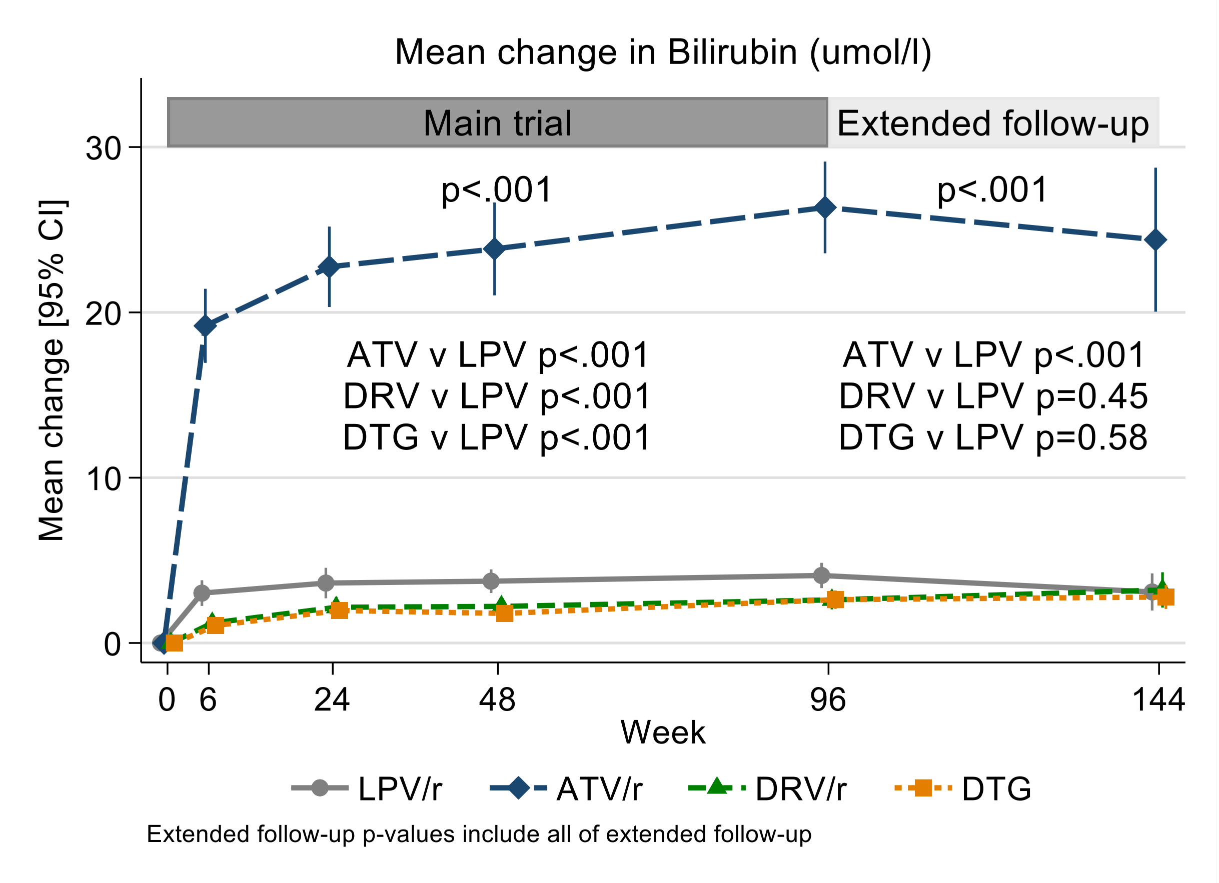

ATV/r denotes ritonavir-boosted atazanavir, DRV/r ritonavir-boosted darunavir, DTG dolutegravir and LPV/r ritonavir-boosted lopinavir

**Figure S8: Change in creatinine clearance**

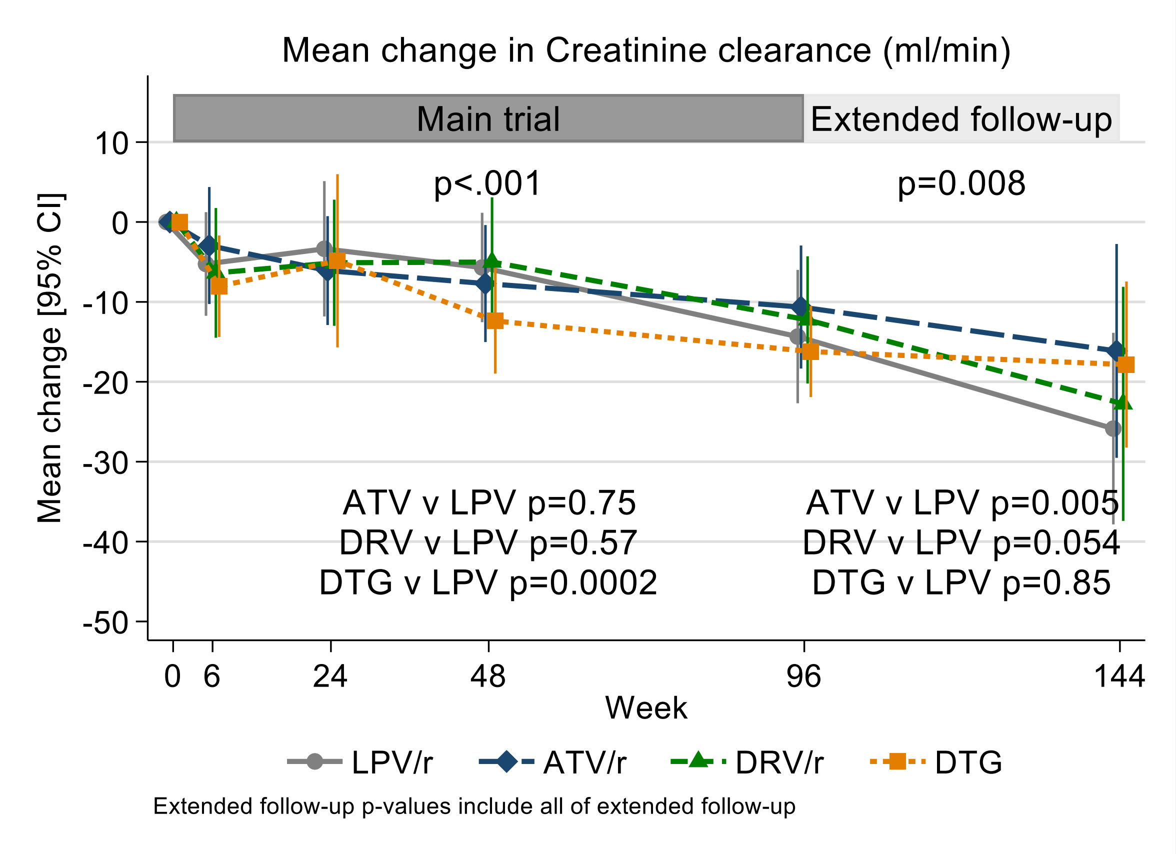

ATV/r denotes ritonavir-boosted atazanavir, DRV/r ritonavir-boosted darunavir, DTG dolutegravir and LPV/r ritonavir-boosted lopinavir

**Figure S9: Change in (a) total, (b) HDL and (c) LDL cholesterol and (d) triglycerides**

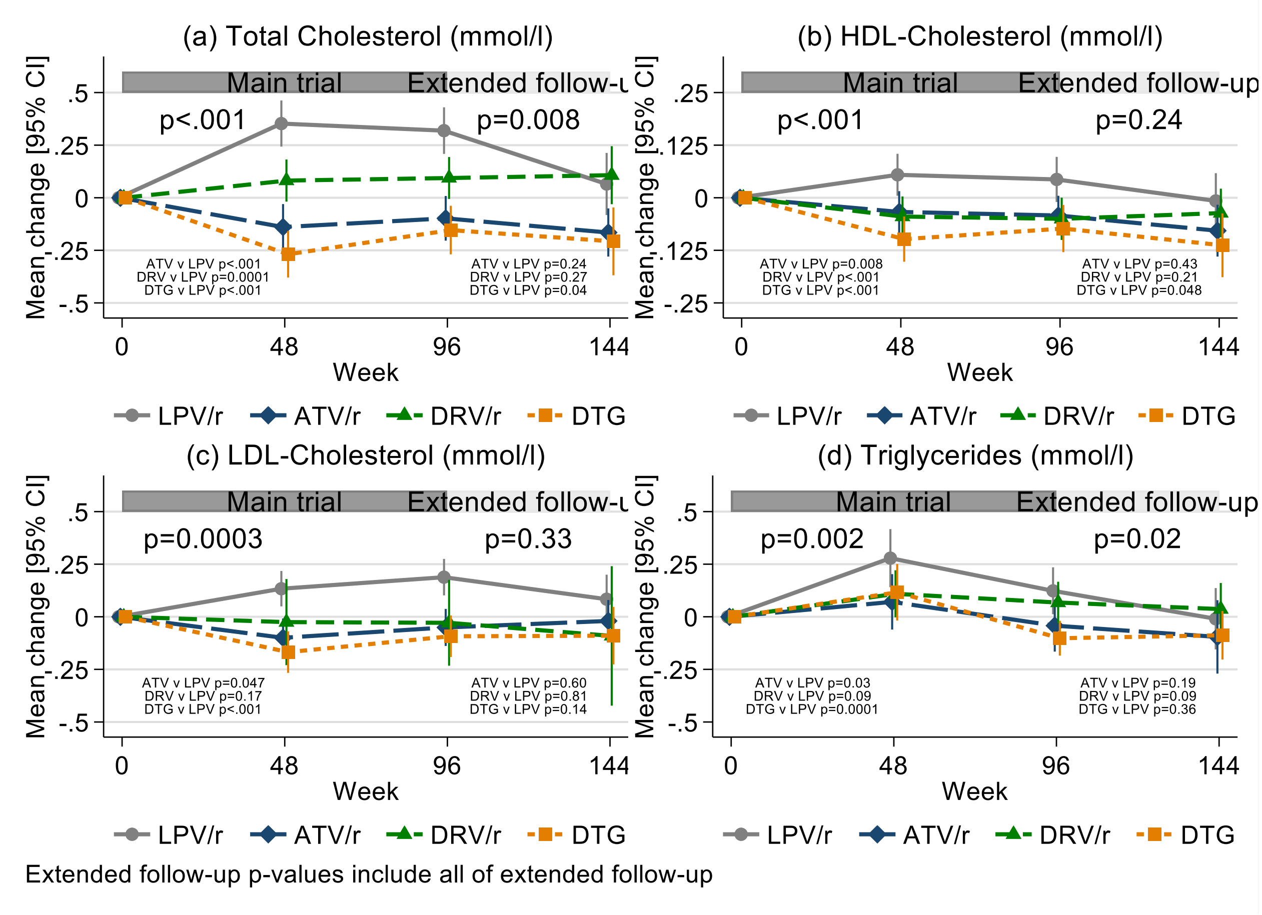

ATV/r denotes ritonavir-boosted atazanavir, DRV/r ritonavir-boosted darunavir, DTG dolutegravir, HDL high-density lipoprotein, LDL low-density lipoprotein and LPV/r ritonavir-boosted lopinavir

**Figure S10: Change in (a) lumbar total and (b) total body less head (i) bone mineral content, (ii) bone mineral density and (iii) bone mineral density Z-score**

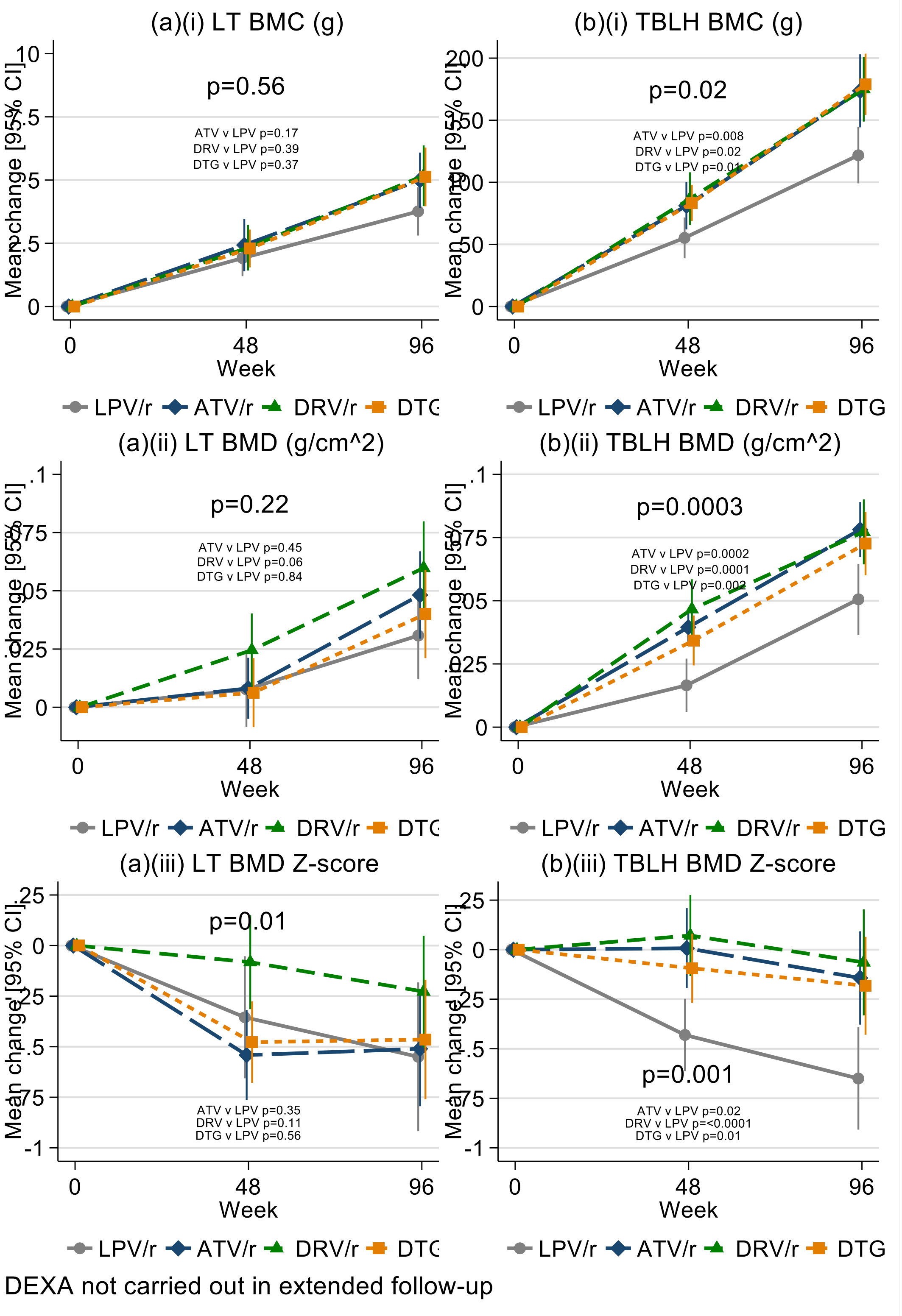

ATV/r denotes ritonavir-boosted atazanavir, BMC bone mineral content, BMD bone mineral density, DRV/r ritonavir-boosted darunavir, DTG dolutegravir, LPV/r ritonavir-boosted lopinavir, LT lumbar total and TBLH total body less head

**Panel S1. The effect of the COVID-19 pandemic**

At the end of March 2020, the trial sponsor and the national authorities made the decision to halt recruitment in all sites due to the COVID-19 pandemic. All enrolled participants continued to be supplied with trial medication and were followed up either at the trial clinic, at home or via phone calls, depending on the level of local lockdown. This had cost and resource implications for the sites, including additional transport and personal protective equipment (PPE). From June 2020, sites restarted recruitment following review of national guidelines and local mitigation plans. The visit window allowed was increased during periods of lockdown or travel/transport restrictions to allow safety and endpoint tests to be conducted. COVID-19 specific protocol deviations were reviewed regularly, and the impact assessed. A manual of operations as well as COVID-19 risk management plans for each site were developed to guide these processes.

Delays in recruitment that arose from this temporary pause in enrolment and the challenges posed by the implementation of 2019 WHO recommendations, which required drug optimization to dolutegravir (DTG) based regimens for all children on first and second-line ART. The independent data monitoring committee (IDMC) and trial steering committee (TSC) agreed to the proposal by trial management group (TMG) that the sample size could be reduced to 920 from 1000 whilst retaining statistical power. This was possible due to the very small loss to follow up (0.5%).
