## Supplemental File 2 for "CHAPAS-4 trial: second-line anchor drugs for children with HIV in Africa"

**Cost-Effectiveness Analysis for Four Anchor Drugs in the CHAPAS-4 Trial**

**Contents**

Methods………………………………………………………………………………………………………………..1

Results………………………………………………………………………………………………………………….1

Table CES1: Costs of antiretroviral drugs……………………………………………………………………..3

Table CES2: Costs of resources used……………………………………………………………………………5

Table CES3. Resource use by anchor drug randomization groups ………………………………..6

Table CES4. Costs and QALYs by anchor drug randomization groups …………………………………7

Table CES5. Costs and Incremental Costs of Different Comparators……………………………..8

Table CES6. Estimated total costs, total QALYs, ICERs and INHBs of different comparators……….9

**Methods**

The economic evaluation assessed the cost-effectiveness over the 96-week trial period of the four anchor drugs in the CHAPAS-4 trial. Health was measured by quality adjusted life years (QALYs) was calculated using the EQ-5D-3L instrument and the area-under-the-curve approach. Total costs were measured from a health care perspective and included antiretroviral (ART) drugs costs, clinic visits costs and hospital stays costs. A discount rate of 3% was applied to costs and QALYs incurred for participants in their second year of the study. All costs were measured in 2022 US$. Costs, QALYs and cost-effectiveness were assessed for the following five comparisons:

(1) dolutegravir (DTG) vs. ritonavir-boosted atazanavir (ATV/r) or ritonavir-boosted lopinavir (LPV/r); (2) ritonavir-boosted darunavir (DRV/r) vs. ATV/r or LPV/r; (3) ATV/r vs. LPV/r; (4) DTG vs. ATV/r or LPV/r or DRV/r. (5) DTG vs. ATV/r vs. LPV/r vs. DRV/r.

Generalized linear models with a gamma distribution and an identity link function were used to estimate the mean total costs and incremental costs and ordinary least squares were used to estimate the mean QALYs and the incremental QALYs. The following stratification factors were controlled for in all regression models: the six sites and the nucleos(t)ide reverse transcriptase inhibitor (NRTI) backbone drugs they failed on in the first line treatment. The baseline EQ5D indices were also controlled for in estimating total and incremental QALYs. Probabilistic sensitivity analyses were conducted to estimate the probability of each intervention being least costly (in cost analysis) or most cost-effective (in cost-effectiveness analysis).

**Results**

Unit costs for the analysis are reported in Tables CES1 and CES2. Resource use and discounted total costs and total QALYs can be found in Table CES3 and CES4.

QALYs were not significantly different across trial arms and there were no systematic trends between arms suggesting any small observed differences were the result of random chance. Therefore, the main health economics analysis focused on assessing the differences in costs between trial arms for the 96-week trial period.

Table CES5 presents mean total costs and incremental costs for each of the comparisons. The four-way comparisons of DTG vs. LPV/r vs. ATV/r vs. DRV/r (comparison 5) showed that DTG was the least costly, and DRV/r most costly. The probability of DTG being cost-saving compared to the other three anchor drugs was 100%. Switching to DTG from the next most costly option, ATV/r, generated a large cost saving of $190.77. Other two-way comparisons suggested that ATV/r was cost-saving compared to LPV/r (saving of $17.35). DTG was cost-saving compared to ATV/r or LPV/r (cost saving of $200.49). DRV/r was around $130.10 more costly than ATV/r or LPV/r.

Table CES6 presents the estimated total costs and total QALYs, and incremental cost-effectiveness ratios (ICERs) and incremental net health benefits (INHBs) for each of the comparisons. The four-way comparisons of DTG vs. LPV/r vs. ATV/r vs. DRV/r showed that LPV/r and DRV/r were dominated by ATV/r. Although ATV/r was more effective (0.0093 QALY), it was also more costly ($190.77) than DTG and the ICER of $20,423 per QALY was much higher than the cost-effectiveness thresholds of £500 per QALY considered here. INHBs of ATV/r, LPV/r and DRV/r compared to DTG were all negative using three different cost-effectiveness threshold levels of £100, £300 and £500 per QALY, suggesting that DTG was the cost-effective option among the four second-line drugs compared. Probabilistic sensitivity analyses also showed that ATV/r, LPV/r or DRV/r had 0% of being cost-effective when compared with DTG. Other two-way comparisons suggested that ATV/r was cost-effective compared to LPV/r with a probability of 99.70%, as it is more effective in terms of QALYs (0.0018 QALY) and slightly less costly ($17.35). DRV/r was dominated by ATV/r or LPV/r being less effective (0.0029 QALY) in terms of QALYs and more costly ($130.10).

Table CES1: Costs of antiretroviral drugs

| | | **WHO weight bands** | | | | |
|---|---|---|---|---|---|---|
| | | **14-19.9 kg** | | **20-24.9 kg** | **25-34.9 kg** | **35- kg (adult)** |
| | Daily dose | 25 mg DT | | 50 mg FCT | 50 mg FCT | 50 mg FCT |
| | Tablets taken | 5*5 mg DT | | 1*50 mg FCT | 1*50 mg FCT | 1*50 mg FCT |
| **Dolutegravir** | **Unit cost($)** | **2.5*DTG (10 mg) disp. scored: 0.05** | **half a DTG (50 mg) FCT: 0.07** | **DTG (50 mg): 0.07** | **DTG (50 mg): 0.07** | **DTG (50 mg): 0.07** |
| **(DTG)** | **Daily cost($)** | **0.125** | **0.035** | **0.07** | **0.07** | **0.07** |
| | Daily dose | 200/75[1] mg | | 200/75[1] mg | 300/100 mg | 300/100 mg |
| | Tablets taken | 2*100 mg ATV, 3*25 mg RTV | | 2*100 mg ATV, 3*25 mg RTV | 1*300/100 mg[2] | 1*300/100 mg[2] |
| | **Unit cost($)** | **ATV 200mg capsule: 0.59, RTV (25 mg) heat-stable: 0.10** | | **ATV 200mg capsule: 0.59, RTV (25 mg) heat-stable: 0.10** | **ATV/r (300/100 mg): 0.41**<br>**ATV 300mg capsule:0.61**<br>**RTV (100 mg) heat-stable: 0.10** | **ATV/r (300/100 mg): 0.41**<br>**ATV 300mg capsule:0.61**<br>**RTV (100 mg) heat-stable: 0.10** |
| | **Daily cost($)** | **0.89** | | **0.89** | **0.41** | **0.41** |
| **Atazanavir/ ritonavir (ATV/r)** | **paediatric drug cost as proportion of adult drug cost** | **0.30** | | **0.30** | **0.41** | **0.41** |
| **Lopinavir/ritonavir** | Daily dose | 400/100 mg | | 400/100 mg | 600/150 mg | 800/200 mg |

| | | | | | |
|---|---|---|---|---|---|
| **(LPV/r)** | Tablets taken | 1*200/50 mg[3] (AM) + 1*200/50 mg[3] (PM) | 1*200/50 mg[3] (AM) + 1*200/50 mg[3] (PM) | 2*200/50 mg[3] (AM) + 1*200/50 mg[3] (PM) | 2*200/50 mg[3] (AM) + 2*200/50 mg[3] (PM) |
| | **Unit cost($)** | **LPV/r (200/50 mg): 0.14** | **LPV/r (200/50 mg): 0.14** | **LPV/r (200/50 mg): 0.14** | **LPV/r (200/50 mg): 0.14** |
| | **Daily cost($)** | **0.28** | **0.28** | **0.42** | **0.56** |
| **Darunavir/ritonavir (DRV/r)** | Daily dose | 600/100 mg | 600/100 mg | 800/100 mg | 800/100 mg |
| | Tablets taken | 4*150 mg[4] DRV, 1*100 mg RTV | 4*150 mg[4] DRV, 1*100 mg RTV | 2*400 mg[5] DRV, 1*100 mg RTV | 2*400 mg[5] DRV, 1*100 mg RTV |
| | **Unit cost($)** | **DRV (150 mg): 0.21, RTV (100 mg) heat-stable: 0.10** | **DRV (150 mg): 0.21, RTV (100 mg) heat-stable: 0.10** | **DRV/r (400/50 mg): 0.32** | **DRV/r (400/50 mg): 0.32** |
| | **Daily cost($)** | **0.94** | **0.94** | **0.64** | **0.64** |
| | **paediatric drug cost as proportion of adult drug cost** | **0.48** | **0.48** | **0.64** | **0.64** |

Notes: ART costs were calculated using data on usage from CHAPAS-4 trial and unit costs of different ART drugs from Clinton Health Access Initiative (CHAI) where available (2022). Where data was unavailable, trial sites provided information on costs. In some cases, we used proportions of the published adult drug costs for the costs of their paediatric drugs (e.g. 200/75 mg ATV/r, 600/100 mg DRV/r).

1. if required 100mg RTV may be used to boost ATV for the 14-19.9kg and 20-24.9kg weight-bands;

2. alternatively three 100mg ATV and four 25mg RTV;

3. double dose of tablets if using 100/25mg, so 2+2 for 14-19.9kg etc;

4. alternatively one 600mg tablet can be used which can be cut; or 8x75mg tablets if available.

DT: dispersible tablet, FCT: film coated tablet, DTG: dolutegravir, ATV/r: ritonavir-boosted atazanavir, LPV/r: ritonavir-boosted lopinavir, DRV/r: ritonavir-boosted darunavir, RTV: ritonavir

Tablets taken: once daily if not stated, or twice daily (morning **AM** and afternoon **PM**)

Table CES2: Costs of resources used

| | Uganda | | Zambia | | Zimbabwe | |
|---|---|---|---|---|---|---|
| | 2022 price in US$ (adjusted for local inflation) | Source | 2022 price in US$ (adjusted for local inflation) | Source | 2022 price in US$ (adjusted for Zambia's inflation as a proxy) | Source |
| Hospital overnight: Teaching hospital | 4.81 | WHO Choice (2011) | 8.83 | WHO Choice (2011) | 3.93 | WHO Choice (2011) |
| Outpatient attendances: Health Centre (no beds) | 0.97 | | 1.78 | | 0.75 | |
| Outpatient attendances: Secondary-level hospital | 1.42 | | 2.60 | | 1.09 | |

Notes: 1. Unit costs of outpatient attendances in a secondary-level hospital (the highest level) were used to cost scheduled and unscheduled visits, unit costs of hospital overnight in a teaching hospital (the highest level) were used to cost hospital stays, and unit costs of outpatient attendances in a health centre (no beds) were used to calculated the costs of visiting a local clinic or healthcare workers; 2. All unit costs were checked with the CHAPAS-4 trial sites to confirm that they were reasonable estimates of the actual costs; 3. Unit cost information collected before 2022 were adjusted for local inflation to get the 2022 price (the unit cost was reported in US dollar in WHO Choice (2011).

Table CES3. Resource use by anchor drug randomization groups

| Anchor drug Randomization groups | LPV/r | | ATV/r | | DRV/r | | DTG | |
|---|---|---|---|---|---|---|---|---|
| | Mean | SD | Mean | SD | Mean | SD | Mean | SD |
| *ART use* | | | | | | | | |
| Average duration of four anchor drugs (days) | 660.28 | 68.79 | 666.78 | 38.01 | 667.38 | 39.07 | 662.17 | 60.21 |
| Average duration of NRTI backbone drugs (days) | 656.33 | 77.67 | 665.57 | 34.37 | 666.72 | 39.78 | 662.39 | 60.26 |
| SOC duration (days) | 641.40 | 119.18 | 660.5 | 48.46 | 658.66 | 72.93 | 668.15 | 23.28 |
| As % of the 96-week trial on SoC | 0.9545 | 0.1773 | 0.9829 | 0.0721 | 0.9801 | 0.1085 | 0.9943 | 0.0346 |
| N (as % of the 919 children) | 115 | 12.51% | 115 | 12.51% | 114 | 12.40% | 117 | 12.73% |
| TAF/FTC duration (days) | 658.21 | 71.59 | 669.66 | 9.33 | 670.37 | 4.22 | 656.38 | 82.58 |
| As % of the 96-week trial on TAF/ FTC | 0.9795 | 0.1065 | 0.9965 | 0.0139 | 0.9976 | 0.0063 | 0.9767 | 0.1229 |
| N (as % of the 919 children) | 112 | 12.19% | 116 | 12.62% | 118 | 12.84% | 112 | 12.19% |
| Average duration of other ART drugs (days) | 2.69 | 40.49 | 2.91 | 44.21 | 0.48 | 7.35 | 0.00 | 0.00 |
| *Other health care use* | | | | | | | | |
| Scheduled clinic visits | 10.72 | 0.96 | 10.84 | 0.56 | 10.83 | 0.73 | 10.78 | 0.80 |
| Unscheduled clinic visits | 1.22 | 2.10 | 1.29 | 2.17 | 0.99 | 1.75 | 0.87 | 2.24 |
| Hospital stays (days) | 0.19 | 1.13 | 0.16 | 1.29 | 0.28 | 1.80 | 0.24 | 3.14 |
| Visits to a local clinic or healthcare workers | 0.22 | 0.63 | 0.25 | 1.42 | 0.20 | 0.60 | 0.22 | 0.72 |
| N (as % of the 919 children) | 227 | (24.70%) | 231 | (25.14%) | 232 | (25.24%) | 229 | (24.92%) |

Notes: 1. The days in four anchor drugs were calculated conditional on patients being allocated to the specific anchor drugs at randomization; 2. We only calculated the resource use when patients were on second-line treatment; 3. Anchor drugs included LPV/r, ATV/r, DRV/r, DTG; 4. NRTI backbone drugs included TAF/FTC and SOC (ABC/3TC or ZDV/3TC); 5. Other ART drugs included TDF/3TC/DTG (300/300/50 mg) and TDF/3TC. DTG: dolutegravir, ATV/r: ritonavir-boosted atazanavir, LPV/r: ritonavir-boosted lopinavir, DRV/r: ritonavir-boosted darunavir, ART: antiretroviral therapy, NRTI: nucleos(t)ide revirse transcriptase inhibitor, SD: standard deviation, SOC: stand-of-care, TAF: tenofovir alafenamide, FTC: emtricitabine, ABC: abacavir, 3TC: lamivudine.

Table CES4. Costs and QALYs by anchor drug randomization groups

| Anchor Drug Randomization Groups | LPV/r | | ATV/r | | DRV/r | | DTG | |
|---|---|---|---|---|---|---|---|---|
| Costs | Mean | SD | Mean | SD | Mean | SD | Mean | SD |
| *ART costs* | | | | | | | | |
| Anchor drugs costs | 269.87 | 77.25 | 247.13 | 36.81 | 390.94 | 57.15 | 51.84 | 15.57 |
| NRTI backbone drugs costs | 117.76 | 37.03 | 120.16 | 34.31 | 118.87 | 33.59 | 122.14 | 33.08 |
| TAF/FTC cost | 48.07 | 50.68 | 48.90 | 50.76 | 50.00 | 51.02 | 48.69 | 51.98 |
| SOC cost | 68.11 | 72.88 | 69.65 | 73.46 | 67.28 | 72.40 | 71.80 | 73.40 |
| Other ART drug costs | 0.30 | 4.45 | 0.30 | 4.51 | 0.06 | 0.96 | 0.00 | 0.00 |
| **Total ART costs** | **382.69** | **96.45** | **362.62** | **56.17** | **502.98** | **78.33** | **171.67** | **34.55** |
| *Other health care costs* | | | | | | | | |
| Scheduled visits costs | 14.88 | 5.50 | 15.06 | 5.42 | 15.02 | 5.56 | 15.11 | 5.65 |
| Unscheduled visits costs | 1.61 | 2.67 | 1.70 | 2.87 | 1.30 | 2.16 | 1.12 | 2.61 |
| Hospital stay costs | 1.31 | 7.83 | 0.69 | 5.30 | 1.42 | 9.32 | 1.08 | 12.85 |
| Cost of visiting a local clinic or healthcare workers | 0.27 | 0.84 | 0.24 | 1.13 | 0.27 | 0.84 | 0.29 | 0.94 |
| **Total other health care costs** | **18.06** | **11.22** | **17.69** | **9.31** | **18.00** | **11.90** | **17.59** | **15.86** |
| **Total health care costs** | **400.73** | **97.30** | **380.31** | **56.02** | **520.81** | **79.06** | **189.26** | **38.88** |
| **QALYs** | **1.8047** | **0.0271** | **1.8045** | **0.0395** | **1.8018** | **0.0304** | **1.7945** | **0.1168** |
| N (as % of the 919 children) | 227 | (24.70%) | 231 | (25.14%) | 232 | (25.24%) | 229 | (24.92%) |

Notes: 1. All the costs were in 2022 US dollar; 2. The discount factor for costs and QALYs occurring in the second year of the study was 3% per annum; 3. Anchor drugs included LPV/r, ATV/r, DRV/r, DTG; 4. NRTI backbone drugs included TAF/FTC, ABC/3TC, ZDV/3TC; 5. Other ART drugs included TDF/3TC/DTG (300/300/50 mg) and TDF/3TC; 6. QALYs were captured by EQ-5D index over 96 weeks using area-under-the-curve approach. Only EQ-5D indices reported in the scheduled visits were used to calculate QALYs. Missing baseline indices were imputed using the sample average index, while other missing indices were imputed using the mean of the indices before and after the missing index of the same individual. DTG: dolutegravir, ATV/r: ritonavir-boosted atazanavir, LPV/r: ritonavir-boosted lopinavir, DRV/r: ritonavir-boosted darunavir, ART: antiretroviral therapy, NRTI: nucleos(t)ide revirse transcriptase inhibitor, SD: standard deviation, SOC: stand-of-care, TAF: tenofovir alafenamide, FTC: emtricitabine, ABC: abacavir, 3TC: lamivudine, QALY: quality-adjusted life year,

Table CES5. Costs and Incremental Costs of Different Comparators

| Comparators | Costs | Incremental cost | | | | Probability of being least costly |
|---|---|---|---|---|---|---|
| | mean | mean | SE | 95% CI | | |
| (1) ATV/r or LPV/r | 389.89 | | | | | 0% |
| DTG | 189.40 | -200.49 | 4.03 | -208.39 | -192.58 | 100% |
| N=687 | | | | | | |
| (2) ATV/r or LPV/r | 390.41 | | | | | 100% |
| DRV/r | 520.50 | 130.10 | 6.63 | 117.10 | 143.10 | 0% |
| N=690 | | | | | | |
| (3) LPV/r | 399.07 | | | | | 0.4% |
| ATV/r | 381.72 | -17.35 | 6.69 | -30.45 | -4.25 | 99.6% |
| N=458 | | | | | | |
| (4) ATV/r or LPV/r or DRV/r | 433.66 | | | | | 0% |
| DTG | 189.56 | -244.09 | 4.34 | -252.60 | -235.58 | 100% |
| N=919 | | | | | | |
| Comparators | Costs | incremental cost compared to the next lowest cost | | | | Probability of being least costly |
| | mean | mean | SE | 95% CI | | |
| (5) DTG | 189.58 | | | | | 100% |
| ATV/r | 380.35 | 190.77 | 4.76 | 181.45 | 200.10 | 0% |
| LPV/r | 399.20 | 18.85 | 6.22 | 6.67 | 31.03 | 0% |
| DRV/r | 520.41 | 121.21 | 7.38 | 106.73 | 135.68 | 0% |
| N=919 | | | | | | |

Notes: 1. All the costs were in 2022 US dollar and a discount rate of 3% per annum was applied to costs incurred for participants in their second year of the study; 2. The model controlled for stratification factors of the six sites and the NRTI backbone drugs they failed on in the first line treatment; 3. Probabilistic sensitivity analyses were conducted to estimate the probability of each intervention being least costly using the threshold of $500 per QALY implied by the decisions that had been previously made on the ART drugs for HIV; 4. There were five comparators of different anchor drugs: (1) DTG vs. ATV/r or LPV/r; (2) DRV/r vs. ATV/r or LPV/r; (3) ATV/r vs. LPV/r; (4) DTG vs. ATV/r or LPV/r or DRV/r. (5) DTG vs. ATV/r vs. LPV/r vs. DRV/r. DTG: dolutegravir, ATV/r: ritonavir-boosted atazanavir, LPV/r: ritonavir-boosted lopinavir, DRV/r: ritonavir-boosted darunavir, ART: antiretroviral therapy, NRTI: nucleos(t)ide revirse transcriptase inhibitor, SE: standard error, CI: confidence interval.

Table CES6. Estimated total costs, total QALYs, ICERs and INHBs of different comparators

| Comparators | Total Cost | Total QALYs | Incremental cost | | | | Incremental QALYs | | | | ICER | INHB (λ=100) | Prob of being CE | INHB (λ=300) | Prob of being CE | INHB (λ=500) | Prob of being CE |
|---|---|---|---|---|---|---|---|---|---|---|---|---|---|---|---|---|---|
| | mean | mean | mean | SE | 95% CI | | mean | SE | 95% CI | | | | | | | | |
| (1) ATV/r or LPV/r | 389.89 | 1.8040 | | | | | | | | | | | 0% | | 0% | | 0% |
| DTG | 189.40 | 1.7957 | -200.49 | 4.03 | -208.39 | -192.58 | -0.0083 | 0.0057 | -0.0194 | 0.0029 | 24171.15 | 1.9950 | 100% | 0.6595 | 100% | 0.3924 | 100% |
| N=687 | | | | | | | | | | | | | | | | | |
| (2) ATV/r or LPV/r | 390.41 | 1.8046 | | | | | | | | | | | 100% | | 100% | | 100% |
| DRV/r | 520.50 | 1.8017 | 130.10 | 6.63 | 117.10 | 143.10 | -0.0029 | 0.0023 | -0.0075 | 0.0017 | Dominated | -1.3010 | 0% | -0.4356 | 0% | -0.2625 | 0% |
| N=690 | | | | | | | | | | | | | | | | | |
| (3) LPV/r | 399.07 | 1.8037 | | | | | | | | | | | 0.30% | | 0.30% | | 0.30% |
| ATV/r | 381.72 | 1.8055 | -17.35 | 6.69 | -30.45 | -4.25 | 0.0018 | 0.0026 | -0.0033 | 0.0068 | -9773.22 | 0.1767 | 99.70% | 0.0601 | 99.70% | 0.0368 | 99.70% |
| N=458 | | | | | | | | | | | | | | | | | |
| (4) ATV/r or LPV/r or DRV/r | 433.66 | 1.8033 | | | | | | | | | | | 0% | | 0% | | 0% |
| DTG | 189.56 | 1.7956 | -244.09 | 4.34 | -252.6 | -235.58 | -0.0077 | 0.0048 | -0.0171 | 0.0017 | 31818.65 | 2.4336 | 100% | 0.8059 | 100% | 0.4803 | 100% |
| N=919 | | | | | | | | | | | | | | | | | |
| Comparators | Total Cost | Total QALYs | Incremental cost compared to the next lowest cost | | | | Incremental QALY compared to the next lowest cost | | | | ICER vs. DTG | INHB (λ=100) vs. DTG | Prob of being CE | INHB (λ=300) vs. DTG | Prob of being CE | INHB (λ=500) vs. DTG | Prob of being CE |
| | mean | mean | mean | SE | 95% CI | | mean | SE | 95% CI | | | | | | | | |
| (5) DTG | 189.58 | 1.7956 | | | | | | | | | | | 100% | | 100% | | 100% |
| ATV/r | 380.35 | 1.8049 | 190.77 | 4.76 | 181.45 | 200.10 | 0.0093 | 0.0059 | -0.0022 | 0.0208 | 20423.25 | -1.9002 | 0% | -0.6272 | 0% | -0.3726 | 0% |
| LPV/r | 399.20 | 1.8035 | 18.85 | 6.22 | 6.67 | 31.03 | -0.0014 | 0.0059 | -0.0130 | 0.0101 | Dominated | -2.0895 | 0% | -0.6912 | 0% | -0.4115 | 0% |
| DRV/r | 520.41 | 1.8014 | 121.21 | 7.38 | 106.73 | 135.68 | -0.0021 | 0.0059 | -0.0136 | 0.0094 | Dominated | -3.3023 | 0% | -1.0968 | 0% | -0.6557 | 0% |
| N=919 | | | | | | | | | | | | | | | | | |

Notes: 1. All the costs are in 2022 US dollar; 2. Generalized linear model with a gamma distribution of the dependent variable and an identity link function was used to estimate the mean total costs and the incremental costs, controlling for stratification factors; 3. Generalized linear model with a gaussian (normal) distribution of the dependent variable and an identity link function (equivalent to ordinary least squares) was used to estimate the mean total QALYs and the incremental QALYs, controlling for stratification factors and baseline EQ5D indices; 4. Probabilistic sensitivity analyses were conducted to estimate the probability of each intervention being cost effective (reported next to the INHBs).; 5. There are five comparators: (1) DTG vs. ATV/r or LPV/r; (2) DRV/r vs. ATV/r or LPV/r; (3) ATV/r vs. LPV/r; (4) DTG vs. ATV/r or LPV/r or DRV/r. (5) DTG vs. ATV/r vs. LPV/r vs. DRV/r; 6. Cost-effectiveness threshold (λ) of $500 per QALY implied by the decisions that had been previously made on the ART drugs for HIV were used to calculate ICER; 7. For scenario analyses, cost-effectiveness thresholds of $100 and $300 per QALY were also used to calculate INHB. DTG:

dolutegravir, ATV/r: ritonavir-boosted atazanavir, LPV/r: ritonavir-boosted lopinavir, DRV/r: ritonavir-boosted darunavir, ART: antiretroviral therapy, NRTI: nucleos(t)ide revirse transcriptase inhibitor, SE: standard error, CI: confidence interval. CE: cost effective, QALY: quality-adjusted life years, ICER: incremental cost-effectiveness ratio**,** INHB: incremental net health benefits.
